# Sparse Polygenic Risk Score Inference with the Spike-and-Slab LASSO

**DOI:** 10.1101/2025.01.28.25321292

**Authors:** Junyi Song, Shadi Zabad, Archer Yang, Simon Gravel, Yue Li

**Affiliations:** School of Computer Science, McGill University; Department of Mathematics and Statistics, McGill University; Department of Human Genetics, McGill University

**Keywords:** Polygenic risk score, Variable selection, GWAS, Sparsity, High-dimensional analysis, Penalized regression, Spike-and-slab LASSO

## Abstract

Large-scale biobanks, with comprehensive phenotypic and genomic data across hundreds of thousands of samples, provide ample opportunities to elucidate the genetics of complex traits and diseases. Consequently, there is a growing demand for robust and scalable methods for disease risk prediction from genotype data. Performing inference in this setting is challenging due to the high-dimensionality of genomic data, especially when coupled with relatively smaller sample sizes. Popular Polygenic Risk Score (PRS) inference methods address this challenge by adopting sparse Bayesian priors or penalized regression techniques, such as the Least Absolute Shrinkage and Selection Operator (*LASSO*). However, the former class of methods are not as scalable and do not produce exact sparsity, while the latter tends to over-shrink large coefficients. In this study, we present *SSLPRS*, a novel PRS method based on the Spike-and-Slab LASSO (SSL) prior, which offers a theoretical bridge between the two frameworks. We extend previous work to derive a coordinate-ascent inference algorithm that operates on GWAS summary statistics, which is orders-of-magnitude more efficient than corresponding individual-level-based implementations. To illustrate the statistical properties of the proposed model, we conducted experiments involving 9 simulation configurations and 9 quantitative phenotypes from the UK Biobank. Our results demonstrate that *SSLPRS* is competitive with state-of-the-art methods in terms of prediction accuracy and exhibits superior variable selection performance, especially in sparse genetic architectures. In simulations, this translates to upwards of 50% improvement in positive predictive value. In analysis of real phenotypes, we show that selected variants are highly enriched for meaningful genomic annotations and have better replication rates in larger meta-analyses.

## Introduction

Polygenic risk scores are emerging as an important tool to quantify the heritable component of complex traits and diseases, with an array of promising clinical applications, including genetic risk stratification and personalized medicine (Torkamani et al., 2018; Lewis and Vassos, 2020). However, inference of polygenic scores from large-scale Genome-wide Association Study (GWAS) data is practically challenging due to two main factors. First, due to privacy concerns, individual-level data is rarely publicly available, and consequently, inference has to be carried out using “GWAS summary statistics” (Pasaniuc and Price, 2017), which are the marginal association statistics per variant, coupled with Linkage-Disequilibrium (LD) matrices that record the pairwise correlations between variants in the dataset. The second factor is the ultra high-dimensional nature of genetic data, with modern biobank initiatives routinely measuring and imputing data for upwards of 20 million genetic markers (Karczewski et al., 2024). Considering that GWAS sample sizes, even for the largest meta-analyses, rarely exceed two million samples (Zhou et al., 2022), this setup calls for robust and scalable regularized or Bayesian regression frameworks.

Previous work has explored various statistical and algorithmic approaches to meet these challenges (Jayasinghe et al., 2024). This includes sparse penalized regression methods, such as `Lassosum` (Mak et al., 2017), as well as a wide array of sparse Bayesian priors, including Horseshoe (`PRScs`) (Ge et al., 2019), Spike-and-slab (`LDPred, VIPRS`) (Privé et al., 2020; Zabad et al., 2023), and sparse mixture priors (`SBayesR, MegaPRS`) (Lloyd-Jones et al., 2019; Zhang et al., 2021). While the overall prediction accuracy of these individual methods is roughly comparable (Pain et al., 2021; Zabad et al., 2023), they can differ substantially in terms of computational resource utilization, the quality of selected variables, and the statistical properties of their inferred effect sizes. On typical GWAS datasets with 1 million genetic variants, the total wallclock time required can range from 15 minutes for optimization-based approaches (*VIPRS*) up to several hours for Markov-Chain Monte-Carlo (MCMC)-based Bayesian methods (`PRScs`) (Zabad et al., 2023). While Bayesian methods can provide accurate and unbiased effect sizes estimates, their coefficients are not truly sparse, which can complicate interpretation and deploying these predictors in some practical settings. On the other hand, traditional penalized regression methods such as the *LASSO* provide sparse effect sizes estimates, but they are known to over-shrink large coefficients, thus introducing a bias (James et al., 2019).

Here, we bridge the gap between sparse Bayesian methods and traditional penalized regression approaches by developing and testing a new PRS model based on the recently-proposed Spike-and-slab LASSO (SSL) prior (Ročková and George, 2018). The *SSL* prior is a mixture of two Laplace densities that together form a non-concave penalty, and through posterior mode estimation, small coefficients are shrunk to exactly zero while preserving larger effect sizes (Ročková and George, 2018; Bai et al., 2021). In addition to fast coordinate-ascent inference algorithms, the *SSL* prior comes with many attractive theoretical properties, and, depending on the setting of its hyperparameters, can smoothly interpolate between the *LASSO* and Spike-and-slab priors (Ročková and George, 2018). Our contributions include the following: **(1)** Deriving a version of the *SSL* inference algorithms that operates on GWAS summary statistics. **(2)** Providing a fast and memory-efficient software implementation that incorporates state-of-the-art techniques (Zabad et al., 2025) for scaling PRS inference to millions of genetic markers. Open source code is available at https://github.com/li-lab-mcgill/penprs. **(3)** Examining the statistical and computational performance of the *SSLPRS* model on both simulated data and real quantitative phenotypes in the UK Biobank (Bycroft et al., 2018). In addition, we provide a new implementation of the *LASSO* model that takes advantage of highly-optimized data structures and algorithms (Zabad et al., 2025), which improves its runtime by more than an order of magnitude compared to the original version (`Lassosum`) (Mak et al., 2017; Zabad et al., 2023).

## Methods

### Overview of SSLPRS

In a sample of *n* individuals with paired genotype and phenotype data of *p* variants, the standard linear model is used to parameterize the dependence of phenotype on the genotype,

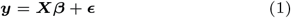

where **y** is an *n×*1 vector of phenotypic measurements for each individual, ***X*** is an *n × p* genotype matrix, ***β*** is a *p ×* 1 vector of effect sizes per variant, and ***ϵ*** is an *n ×* 1 vector representing the residual effects of the phenotype for each individual. We adapted the *SSL* framework (Ročková and George, 2018), originally operating on individual level data, to support GWAS summary statistics for polygenic risk prediction.

The *SSL* prior primarily consists of two Laplace densities, *ψ*(*β* | *λ*) = (*λ/*2)*e*^−*λ*|*β*|^ to model the “spike” and the “slab” respectively. This formulation facilitates the adaptive shrinkage of variants based on their covariance and the associations with the target phenotype:

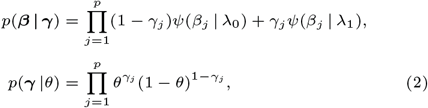

where *θ* ∼ *Beta*(*a, b*).

Typically, the scale parameters are chosen such that *λ*_0_ ≫ *λ*_1_, so *ψ*(· | *λ*_1_) is diffuse, representing a slab with respect to the spiky *ψ*(· | *λ*_0_) that is dense around zero. The Laplace distributions of the prior introduce essentially two *LASSO* components where effect sizes of variants assigned to the “spike” component can be driven to exact sparsity by a strong *λ*_0_ penalty, whereas those in the “slab” component undergo usually minimal shrinkage by a relatively weaker *λ*_1_ penalty. Here, *θ* ∈ (0, 1) denotes the mixing proportion, and the shared *θ* prior in Equation 2 renders the *SSL* penalty non-separable, enabling *SSL* to borrow information across coefficients and adapt to sparsity patterns. (Bai et al., 2021; Ročková and George, 2018). The prior on ***β*** can be parameterized on *θ* by marginalizing out ***γ***:

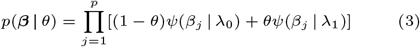

The selection of *λ*_0_ and *λ*_1_, specifically the ratio *λ*_0_*/λ*_1_ places *SSL* on the continuum of the *LASSO* case and the ideal spike-and-slab (Figure 1a). In the case where *λ*_0_ = *λ*_1_ or *λ*_1_ = 0, *SSL* simplifies to the *LASSO l*_1_ penalty (Figure 1b). On the other hand, choosing *λ*_0_ → ∞ gives a theoretical point-mass spike (Ročková and George, 2018).

**Fig. 1.**
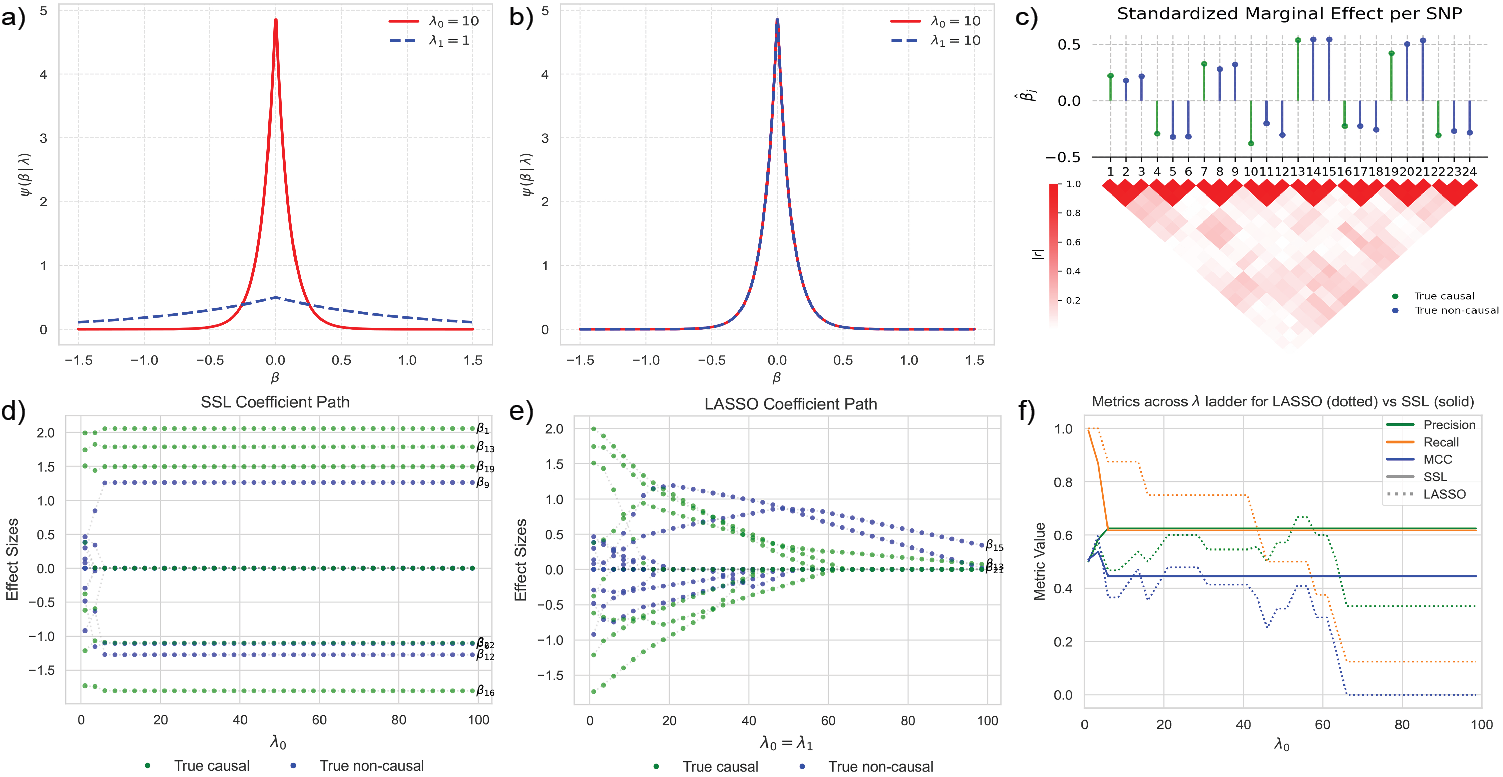
Overview of the statistical properties and inference dynamics of the *SSL* and *LASSO* on simulated genetic data. (a, b) Illustration of the densities *ψ*(*β* | *λ*_0_), *ψ*(*β* | *λ*_1_) of the *SSL* prior with (a) *λ*_0_ = 10 and *λ*_1_ = 1 (b) *λ*_0_ = *λ*_1_ = 10, which reduces to the *LASSO* case. (c) Simulated genetic data with 24 genetic variants with block-diagonal correlation structure. In this setup, 8 of the variants are causal (green) while 16 are non-causal (blue). Marginal effect sizes are shown in the top panel and absolute pairwise correlations between the variants are shown in the bottom panel. (d, e) Effect size estimates of (d) SSL and (e) *LASSO* models are shown along a ladder of increasing *λ*_0_ values. Ground truth causal variants are shown in green, while non-causal ones are shown in blue. Non-zero effect size estimates at the end of *λ* ladder are labeled with their corresponding index. (f) Variable selection metrics, including precision, recall, and Matthews Correlation Coefficient (MCC), for the *LASSO* and *SSL* models across the *λ* ladder.

### SSLPRS inference

(Ročková and George, 2018; Moran et al., 2019) introduced an efficient coordinate ascent Maximum A Posteriori (MAP) algorithm for *SSL*. In this work, we developed an improved framework of the coordinate ascent algorithm with support for GWAS summary statistics, alongside inference optimizations. The objective function is the log posterior, under the assumptions that both the genotype matrix ***X*** and phenotype vector ***y*** have been standardized column-wise for unit variance and zero mean for GWAS summary statistics:

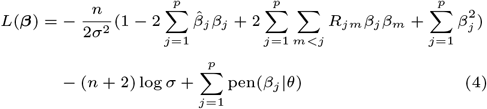

where *N* is the GWAS sample size.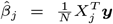 denotes the standardized marginal GWAS effect size for SNP *j* with *X*_*j*_ as the *j*-th column of ***X. R*** = (*R*_*jm*_) defines the *p × p* Linkage Disequilibrium (LD) matrix with 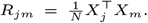 The last term in Equation (4) is the separable spike-and-slab Lasso penalty (Ročková and George, 2018):

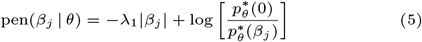

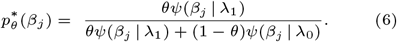

In high-dimensional analysis, the posterior is likely to be multi-modal. The Karush–Kuhn–Tucker (KKT) conditions, coinciding with the standard *LASSO* estimate criteria (Zhao and Yu, 2006), provide only the necessary condition for for ***β***^**∗**^ to be a global mode under the *SSL* prior, expressed as soft-thresholding on *β*_*j*_. Sufficient conditions require the filtering of sub-optimal local modes, achieved through hard-thresholding with Δ (Moran et al., 2019; Ročková and George, 2018; Zhang and Zhang, 2012). Combining these conditions gives

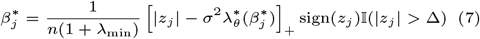

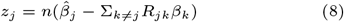

and 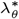 corresponds to the partial derivative of pen(*β*_*j*_ | *θ*) with respect to |*β*_*j*_ | (Ročková and George, 2018), *and is expressed as:*

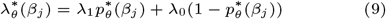

Here, Δ ≡ inf_*t>*0_[*nt/*2 − *σ*^2^pen(*t* |*θ*)*/t*], which can be approximated (Moran et al., 2019) by:

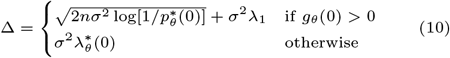

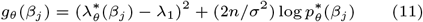

In our initial experiments, we found that fitting *SSLPRS* with sparse LD matrices may encounter numerical instabilities, due to the fact that these matrices lose their positive semi-definite (PSD) property (Zabad et al., 2025). To ensure stability, we introduced *λ*_min_, a quantity derived from spectral properties of the LD matrix, in Eq. 7 to regularize the *β*_*j*_ update and avoid modes aligned with negative eigenvalues (Details in Supplemental Methods S3.5).

The global mode can subsequently be determined through an efficient coordinate ascent algorithm (Algorithm 1), applied to summary statistics, by iteratively updating the model parameters until convergence. By Lemma 4 from (Ročková and George, 2018), *θ*^(*k*)^ can be approximated by

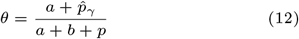

where 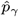 is the number of non-zero coefficients.

In this work, we kept *σ*^2^ constant as a hyperparameter. A variation of the coordinate ascent algorithm is the unknown variance case (Bai et al., 2021), which we present in full for summary statistics in the Supplemental methods S3.3.

### Hyperparameter choices

The default *SSLPRS* model operates on the fixed variance case, where we set *σ*^2^ = 1 based on the assumption that the data is standardized. For hyperparameters *a, b* in the *θ* update, we set *b* = *p* and 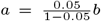 so the beta prior mean is 0.05 (Eq. 2), reflecting an estimate of 5% causality in the dataset. The selection of penalty values *λ*_0_, *λ*_1_ scales with the data through 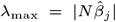, which is the maximum penalty *LASSO* can take before all effect sizes are shrunk to zero (Friedman et al., 2010). To ensure that the slab is sufficiently diffuse, *λ*_1_ is set to *ϵλ*_max_, where *ϵ* = 10^−3^. In the single-fit case, *λ*_0_ is scaled to be *λ*_0_ = 100*λ*_1_ or 10% of *λ*_max_. *λ*_min_ is set to the minimum value for the LD matrix to remain positive semi-definite and numerically stable (Zabad et al., 2025). Details of hyperparameter choices and corresponding ablation analyses can be found in Supplemental Results S4.3.

#### Algorithm 1

SSLPRS Coordinate Ascent

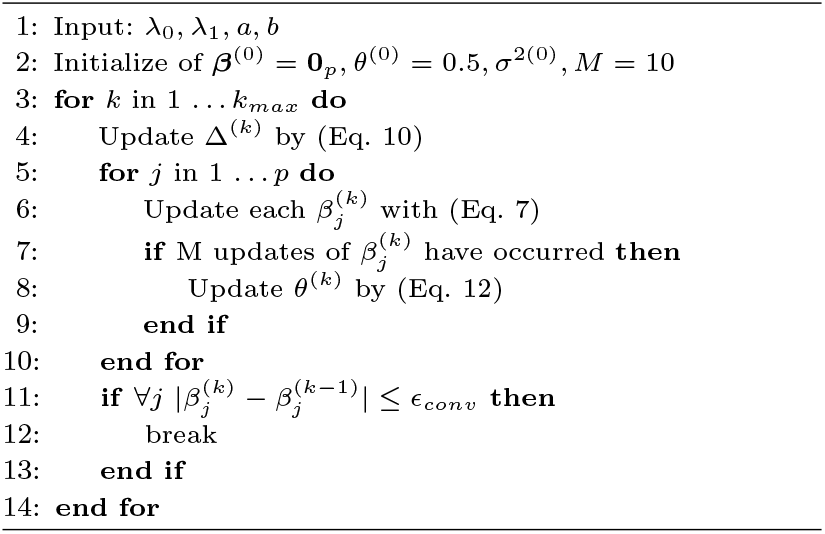

#### Algorithm 2

SSLPRS Dynamic Warm-start.

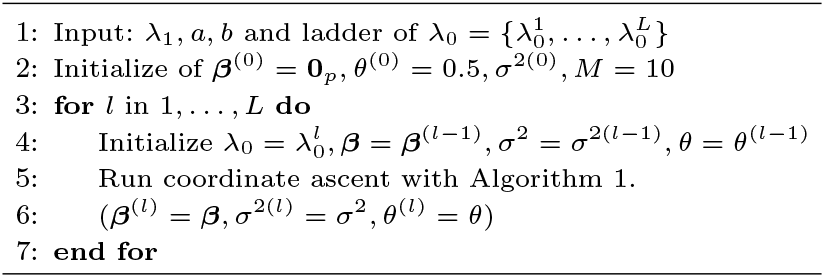

#### Dynamic posterior exploration

Since the choice of hyperparameters *λ*_0_ and *λ*_1_ greatly affects the shrinkage and degree of the spike and slab formulation of *SSLPRS* on the continuum between *LASSO* and the point mass spike and slab, optimizing the performance requires a precise selection of these hyperparameters. (Ročková and George, 2018) detailed a dynamic posterior exploration strategy, where a ladder of gradually increasing *λ*_0_ values 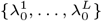 are selected while *λ*_1_ is kept at a sufficiently diffuse value relative to the choices of *λ*_0_ in the ladder. The initial 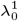 value in the ladder is typically selected such that (*λ*_1_ − *λ*_0_)^2^ *<* 4, which makes the objective (Eq. 4) convex (Bai et al., 2021). The convex solution can be used as a “warm-start” for non-convex problems, which occurs when *λ*_0_ ≫ *λ*_1_. In particular, after fitting each 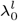 on the ladder with Algorithm 1, the MAP estimation of ***β***, along with model parameters (Δ, *θ, σ*^2^) are used to “warm-start” the fit for 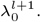 (Algorithm 2.)

Our experiments show that *λ*_max_ becomes the minimum penalty beyond which all negligible coefficients would be set to zero (Friedman et al., 2010), essentially “converging” along the coefficient path of the *λ*_0_ ladder. This also provides a strict range of *λ*_0_ values to search in for hyperparameter tuning. By default, *SSLPRS* performs a warm-start exploration on a 20-step log_2_ scaled *λ*_0_ ladder from *λ*_1_ to *λ*_max_, which does not require grid search.

#### Hyperparameter search

In the grid-search (*SSLPRS-GS*) case, hyperparameter search is conducted independently for each chromosome, where effect size estimates are taken from the best performing *λ*_0_ value of the warm-start ladder on the validation set. Since warm-start is performed to the end of the ladder, *SSLPRS* can be concurrently fit as well. Hyperparameters *a, b* are kept at their default setting of 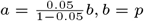 and *σ*^2^ is set fixed to 1.

### Baseline models and their specifications

To assess the relative performance of the *SSLPRS* model on real GWAS data, we compared it to a baseline C+T method *PRSice2* (Choi and O’Reilly, 2019), and three state-of-the-art PRS inference methods: *Lassosum* (Mak et al., 2017), *VIPRS* (Zabad et al., 2023) and *PRS-CS* (Ge et al., 2019). *Lassosum* imposes a single Laplace density, i.e. *l*_1_ penalty, on the effect sizes and performs coordinate ascent on the penalized loglikelihood to estimate the coefficients. *VIPRS* employs a Gaussian Spike-and-slab prior and uses Coordinate Ascent Variational Inference (CAVI) to approximate the posteriors. *PRS-CS* assigns a continuous shrinkage prior and performs Gibbs sampling. Comparisons with *Lassosum* and *VIPRS* are particularly informative, as *SSL* can theoretically interpolate between their respective formulations, thereby highlighting its statistical flexibility and computational properties (Ročková and George, 2018). At the same time, evaluating against *PRSice2* and *PRS-CS* provides a complementary baseline for widely used and state-of-the-art alternatives.

To enable fair comparisons, we included our optimized implementation, *LASSO*, of *Lassosum* (Mak et al., 2017) using optimizations described in Supplemental methods S3.6, as provided in the open source software package `penprs v0.0.1`. By default, the *LASSO* performs coordinate ascent inference using the pathwise algorithm (Friedman et al., 2010), on a grid of 20 values for the penalty hyperparameter *λ*, ranging from *λ*_*max*_ to 10^−3^*λ*_*max*_ on a log_2_ scale. *Lassosum* used a 20 point grid *λ* grid on a log-scale from 0.001 to 0.1. For the *VIPRS* method (Zabad et al., 2023, 2025), we tested the standard EM algorithm that does not require any hyperparameter tuning as well as a grid-search (*VIPRS-GS*) version that tunes the hyperparameter *π*, which corresponds to the prior mean on the proportion of causal variants. We used a 20 point grid for *π* on a log_10_ scale from 1^−*p*^ to 1 − 1^−*p*^, where *p* is the number of variants used during inference. For *PRS-CS*, we used the auto variant where the *φ* hyperparameter is inferred automatically. All models were run with four threads using parallelism when available.

### UK Biobank data

#### Real phenotype analyses

To empirically examine the performance of *SSLPRS* on real complex traits and diseases, we leveraged paired genotype and phenotype data from 337 205 unrelated White British samples in the UK Biobank (Bycroft et al., 2018). We briefly summarize below the data pre-processing and Quality Control (QC) pipeline that was described in detail in earlier reports by the same group (Zabad et al., 2023, 2025), resulting in a set of 1 093 308 high quality SNPs used in subsequent analyses.

The genotype data, which represents the features used for prediction, was extracted by applying standard quality control filters along the sample and variant dimensions. For the complex traits, which represent the target of our predictors, we extracted measurements for nine quantitative phenotypes for the White British samples described earlier. The phenotypes are standing height (HEIGHT), body mass index (BMI), hip circumference (HC), waist circumference (WC), birth weight (BW), Forced Vital Capacity (FVC), Forced Expiratory Volume in the first second (FEV1), High-density Lipoprotein (HDL), and Low-density Lipoprotein (LDL). The detailed pipeline is described in Supplemental Methods S3.1.2.

To facilitate robust analyses of the predictive performance of the *SSLPRS* and baseline models, we performed 5-fold cross-validation per phenotype. In each round, 80% of the data was used for training the models while the remaining 20% are used for testing; the training set was further split into 90% training and 10% validation for models requiring hyperparameter tuning. We performed genome-wide association testing within each split and generated GWAS summary statistics for the training, validation, and test sets. The 5-fold cross-validation GWAS data are available at https://zenodo.org/records/14612130. The association testing within each split was done with `plink2` (Chang et al., 2015).

Finally, we utilized Linkage-Disequilibrium (LD) matrices for European samples to record pairwise correlations between genetic variants published in a recent study (Zabad et al., 2025), where they served as input to PRS inference methods: https://shz9.github.io/viprs/download_ld/. The matrices were estimated from 362 446 unrelated European samples in the UK Biobank (Bycroft et al., 2018).

#### Simulation experiments

To assess the variable selection performance and capabilities of the *SSLPRS* model under controlled conditions, we used the `magenpy v0.1.5` package (Zabad et al., 2023, 2025) to simulate phenotype data for the UKB participants described above according to a variety of genetic architectures. The phenotypes were simulated using heritability values *h*^2^ = {0.1, 0.3, 0.5}, and causal proportion of variants *θ* = {0.01, 0.001, 0.0001}, for a total of 9 simulation settings. Note that the `magenpy` simulator draws effect sizes according to the Gaussian spike-and-slab generative model (Zabad et al., 2023). We’ve also included a mixture of normals simulation in Supplemental Figure S5. For each setting, we simulated five independent replicates, for a total of 45 simulated phenotypes. The `magenpy` simulator outputs the true causal variants and their effect sizes, and this information was used to examine the variable selection accuracy of each method.

To examine predictive performance in the simulation experiments, the UKB samples were split into 70% training, 15% validation, and 15% testing. The validation set was used for hyperparameter tuning of the grid search (GS) models. These sub-cohorts were then used to perform GWAS using `plink2` (Chang et al., 2015) to obtain marginal association statistics.

### Evaluation metrics and criteria

We evaluated the *SSLPRS* and baseline models based on a number of metrics that account for both computational and statistical performance. The statistical evaluation criteria include prediction accuracy on held-out test sets as well as a number of metrics that quantify variable selection accuracy. In the context of our simulation analyses, the latter includes metrics such as precision (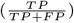), recall (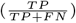), and the Matthews Correlation Coefficient (Ročková and George, 2018) 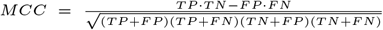 which accounts for all confusion matrix components and thus offers a balanced measure of selection accuracy. Here, *TP* refers to true positives, *TN* true negatives, *FP* false positives, and *FN* false negatives. For Bayesian models like *VIPRS*, we experimented with different Posterior Inclusion Probability (PIP) thresholding for variable selection (Supplemental results S4.3.6) and found that selecting variants with the median rule of PIP *>* 0.5 (Ishwaran and Rao, 2005) worked the best.

To assess the biological significance and replicability of the selected variants, we measure the fold enrichment, defined as

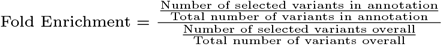

for quantifying the relative concentration of selected variants within a specific annotation compared to their prevalence in the overall background, and the replication rate, which is the proportion of selected variants that is also considered significant in an separate, independent study.

For the grid-search models, we evaluated the prediction performance of hyperparameters in terms of the proportion of variance explained (R-squared). Given a set of inferred coefficients ***β***^∗^, the R-Squared on the validation set can be approximated from GWAS summary statistics via the pseudo-*R*^2^ metric (Mak et al., 2017; Zabad et al., 2023):

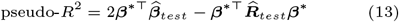

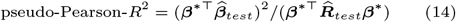

Where 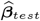 are the standardized marginal GWAS effect sizes from the test set and 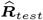 is the corresponding LD matrix. Although pseudo-*R*^2^ offers superior discriminating power for hyperparameter selection, it can be problematic when sparsified LD matrices are used to evaluate external models trained on different reference panels. Thus, we adopted the more robust pseudo-Pearson-*R*^2^ for evaluation on held-out test sets.

In addition to these statistical performance metrics, we examined total wallclock time (in minutes), peak memory utilization (in MB), and inference time (in seconds). Total wallclock time includes the time to load the GWAS and LD data, perform harmonization, load software dependencies, perform inference, and finally conduct cross-validation for model selection (Zabad et al., 2025). Peak memory utilization records the maximum amount of Random Access Memory (RAM) used throughout the lifetime of the program in Megabytes. Finally, inference/fit time records the number of seconds it takes to reach convergence using the coordinate-ascent inference procedure.

## Results

### Simple simulation study

To illustrate the properties of the SSL prior, we designed a simple simulation experiment similar to the setup presented by (Bai et al., 2021), with a sample of *n* = 50 for *p* = 24 SNPs on highly correlated blocks, visually illustrated in Figure 1c. Details can be found in Supplemental Methods S3.1.1.

Fitting *SSLPRS* on this simulated data with an increasing *λ*_0_ ladder through warm-start reveals the model’s ability to simultaneously perform variable selection and stable effect size estimation, i.e. the effects of selected variants are held steady and the rest are shrunk to zero (Figure 1d). Despite the tight correlation structure, *SSLPRS* performs well across the ladder in variable selection in terms of MCC. (Figure 1f). In contrast, the *LASSO* over-shrinks effects with increasing penalization(Figure 1e), which in turn also leads to unstable and degraded variable selection performance across the ladder.

### Simulation of UK Biobank data

To evaluate variable selection and predictive capabilities under diverse genetic architectures, we conducted a 5-replicate analysis on each of the 9 simulation settings(Section 2.5.2). In these analyses, the base *SSLPRS* model is compared to *SSLPRS-GS* and the baseline models. Warm-starting *SSLPRS* to the ladder’s end substantially improves precision, with a modest trade-off in recall, yielding a net gain in MCC in most cases. This behavior is detailed in Supplemental Results S4.1.

The 5-replicate results are shown in Figure 2, with exact numerical values in Supplemental Table S2. Across all settings, *SSLPRS* exhibits lower recall but substantially higher precision compared to *LASSO* and *SSLPRS-GS*. This difference arises because *SSLPRS* selects variants at the end of the *λ*_0_ ladder, resulting in fewer but more confidently identified causal variants. In contrast, the grid-search based *LASSO* and *SSLPRS-GS* typically select lower penalty values, leading to broader but less precise variant selection. Overall, *SSLPRS* performs a balanced variant selection, resulting in a superior MCC performance across most settings. Though, in the less sparse settings (*θ* = 0.01), causal variants are more numerous but have smaller effect sizes, especially under low heritability. Although, *SSLPRS* has slightly lower but still competitive MCC compared to the best performing model in these challenging polygenic settings, it still achieves high precision among the models.

**Fig. 2.**
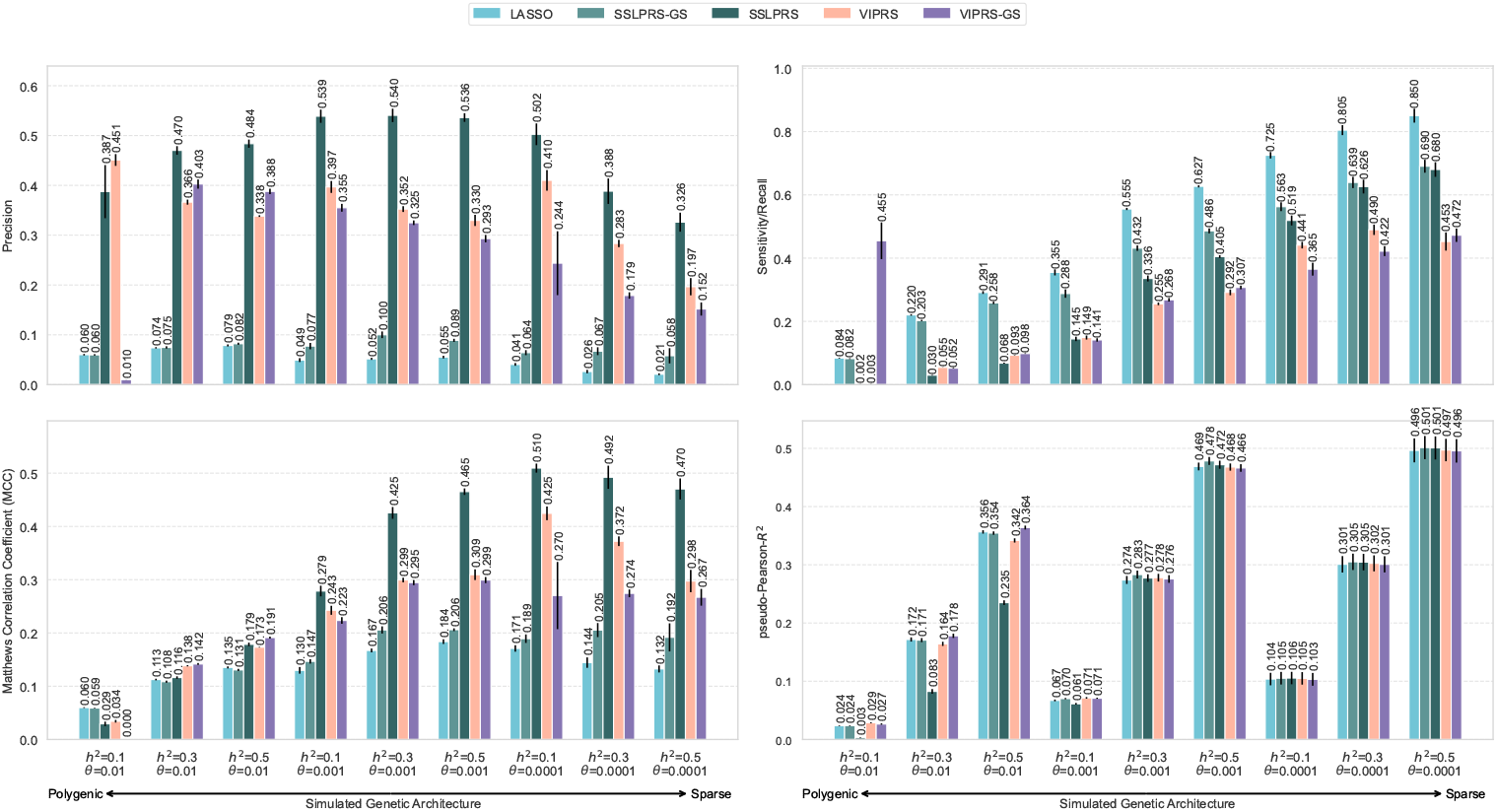
Variable selection and predictive performance of summary statistics-based PRS methods on simulated UK Biobank phenotypes. Performance is evaluated using four metrics: Precision (top-left), Recall/sensitivity (top-right), Matthews correlation coefficient (MCC, bottom-left), and pseudo-Pearson-*R*^2^ (prediction accuracy, bottom-right) on held-out test set. The simulation configurations span three heritability settings, *h*^2^ = {0.1, 0.3, 0.5}, and three proportions of causal variants, *θ* = {0.01, 0.001, 0.0001}. Five summary statistics-based PRS methods are included in this figure: *LASSO*, our proposed *SSLPRS* and *SSLPRS-GS* (grid search), as well as *VIPRS* and *VIPRS-GS* (grid search). Bars represent the mean value over 5 independent replicates. The black vertical lines denote the standard error across the replicates. For the variable selection metrics, the median rule was applied for the *VIPRS* models, where variants with Posterior Inclusion Probability (PIP) *>* 0.5 were considered as selected.

In terms of predictive performance, Figure 2 reveals that in the polygenic (*θ* = 0.01) scenario, *SSLPRS-GS* is more robust in terms of predictive performance compared to *SSLPRS. SSLPRS-GS* achieves pseudo-Pearson-*R*^2^ scores that are comparable to or exceed those of the baseline models. Notably, *SSLPRS-GS* is either on-par or outperforms *LASSO* in all settings. While *SSLPRS* performs well in variable selection, incorporating grid search may further enhance predictive performance, particularly in complex genetic architectures where optimization of pseudo-Pearson-*R*^2^ should be optimized.

### Real phenotype analysis in the UK Biobank

To examine the predictive performance of the *SSLPRS* model on real data, we conducted a 5-fold cross-validation analysis on 9 quantitative phenotypes from the UK Biobank (Bycroft et al., 2018). As this analysis focuses on predictive performance, we compared *SSLPRS-GS*, for the best *SSLPRS* performance, to the main baseline models (Section 2.4). Our results show that across all phenotypes examined, *SSLPRS-GS* shows competitive prediction accuracy compared to the baseline models (Figure 3). Small but significant differences between the models are observed for some phenotypes. For instance, in standing height, models that use grid search to tune hyperparameters outperform *VIPRS* and *PRS-CS* by up to 10%, and in LDL cholesterol, improvements of up to 16% are shown when compared to *PRS-CS*. Across phenotypes, *SSLPRS-GS* matches or outperforms *LASSO*, with advantages in traits such as LDL cholesterol (Figure 3). Interestingly, LDL is a trait known to have a sparse genetic architecture driven by large effect variants (Graham et al., 2021). This may be a case where bias from excessive shrinkage hurts the performance of the *LASSO*, whereas *SSLPRS-GS* benefits from high-quality sparse effect size estimates in a well-selected variant set. Earlier simulations (Figure 2) similarly demonstrated *SSLPRS-GS* performs best under sparse genetic architectures, suggesting that real traits with comparable configurations may yield substantial predictive gains. It’s worth noting that although *LASSO* and *Lassosum* share the same prior formulation, differences in LD scaling, where *Lassosum* tends to an elastic-net like solution, along with other minor variations, can lead to trait-specific performance, with *LASSO* performing better for traits like HDL and LDL, and *Lassosum* for traits like BMI and FEV1 (Details in Supplemental Results S4.2). In addition, we assessed model predictive performance transferability to minority populations (Supplemental Results S4.4).

**Fig. 3.**
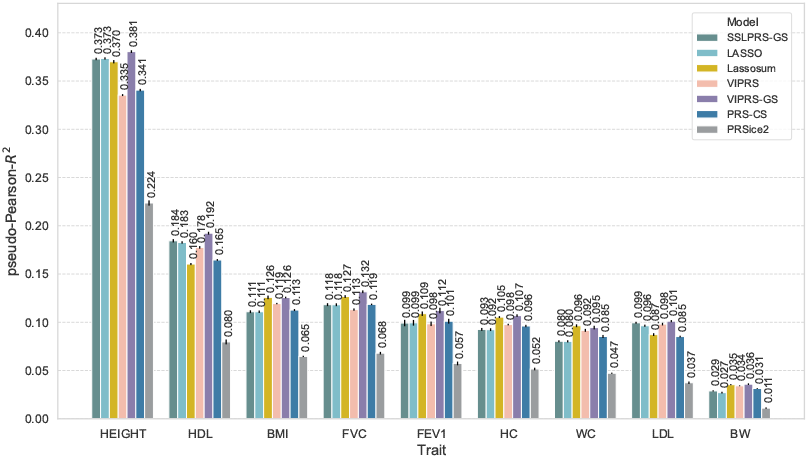
Predictive performance of summary statistics-based PRS methods on real quantitative phenotypes in the UK Biobank. Prediction accuracy (pseudo-Pearson-*R*^2^) on held-out test sets in an analysis of 9 quantitative phenotypes: standing height (HEIGHT), high-density lipoprotein (HDL), body mass index (BMI), forced vital capacity (FVC), forced expiratory volume in 1 s (FEV1), hip circumference (HC), waist circumference (WC), low-density lipoprotein (LDL), and birth weight (BW). The bars represent the average model performance based on 5-fold cross-validation, with black vertical lines indicating the associated standard errors. Six summary statistics-based PRS models are shown with different colors: our proposed *SSLPRS-GS* (grid search), *LASSO, Lassosum, VIPRS, VIPRS-GS* (grid search), PRS-CS (auto), PRSice2.

### Enrichment and replication of selected variants

To assess the biological relevance of selected variants, we evaluated the functional enrichment across 39 genomic annotations (Finucane et al., 2015) and replicability in large-scale GWAS studies. Figure 4a reveals that top performing models in variable selection scenarios, *SSLPRS* and *VIPRS*, exhibit substantial fold-enrichment in some of the biological annotations typically used in partitioning trait heritability (Finucane et al., 2015). Notably, the variants are most enriched in non-synonymous sites, altering amino acid sequence and thus protein function in lipid-regulating genes, and regulatory elements such as promoters and enhancers, underscoring their functional relevance for traits such as HDL and LDL.

**Fig. 4.**
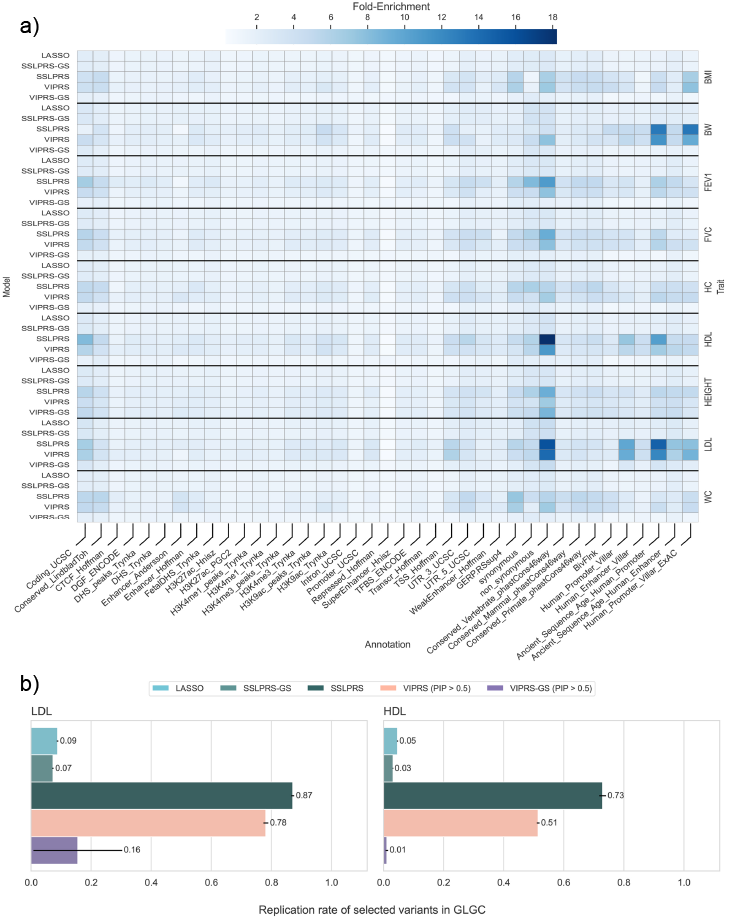
Functional enrichment and biological relevance of selected variants in real complex trait analyses. (a) Heatmap of fold enrichment, defined as the ratio between the proportion of selected SNPs in a given annotation and the background proportion of all SNPs in that annotation. Enrichment is shown across 39 functional annotations (x-axis), stratified by model (left y-axis) and trait (right y-axis). (b) Replication rate of selected variants during training on UK Biobank data for high- and low-density lipoprotein (HDL and LDL) phenotypes in the Global Lipids Genetics Consortium (GLGC) meta-analysis. Replication is defined in terms of the variants being significantly associated with the phenotype (p-value *<* 5 *×* 10^−8^) in the GLGC. Bars and black horizontal bars represent mean replication rate and standard error across the 5-fold cross-validation analyses respectively. 5 summary statistics-based PRS methods are included: *LASSO*, our proposed *SSLPRS* and *SSLPRS-GS* (grid search), as well as *VIPRS* and *VIPRS-GS* (grid search).

To see if selected variants are replicated in large-scale GWAS analyses, we examined summary statistics provided by the Global Lipids Genetics Consortium (GLGC) (Graham et al., 2021), with samples sizes exceeding 1.6 million. We assumed that significant associations reported by that study (p-values < 5 × 10^−8^) as our reference and examined the number of selected variants from the UK Biobank data that are replicated in that larger study. Figure 4b reveals that a substantial proportion of selected variants attain genome-wide significance in the GLGC meta-analysis; *SSLPRS* exhibits the highest replication rate, followed by *VIPRS* in both the LDL and HDL traits. The number of selected, significant, and replicated variants for each model across the 5-folds can be found in Supplemental Table S3. Together, these results indicate that SSLPRS yields a high-quality, sparse set of biologically relevant variants.

### Scalability and computational performance

To assess the computational performance of *SSLPRS* with the optimizations detailed in Supplemental Methods S3.6, we compared our summary statistics implementation of *SSL* to the individual-level R-based SSLASSO (Ročková and George, 2018), benchmarked on an identical chromosome 22 dataset from the UK Biobank standing-height data. SSLPRS achieved a multi-order improvement compared to SSLASSO. Specifically, across settings, *SSLPRS* had an average inference time of 5.3s and a peak memory usage of 73MB, compared to 22.4 min and 41 00 MB for SSLASSO (Details in Supplemental Results S4.6).

## Discussion

In this work, we presented the novel PRS method *SSLPRS* for sparse and accurate prediction of complex traits from GWAS summary data. Our experiments on real and simulated traits show that *SSLPRS* combines competitive prediction accuracy with strong variable selection performance for effective and interpretable PRS construction, especially for sparse genetic architectures. In particular, we emphasized variable selection because the prior, while not perfect, proves more accurate than existing methods in this aspect. This ability is valuable for downstream applications where efficiently obtaining a limited subset of variants is important, and for pipelines that rely on heuristic selection strategies such as P+T or C+T, *SSLPRS* could provide both improved accuracy and efficiency. Through its dynamic posterior exploration with the mode-targeted MAP coordinate ascent algorithm, *SSLPRS* ‘s performance is complemented by its ability to produce truly sparse effect size estimates — retaining a high quality set of biologically significant variants without excessive shrinkage. In addition *SSLPRS* comes with fast, memory-efficient, and highly scalable algorithms for genome-wide inference, available in penprs v0.0.1. Further extensions could explore posterior mean estimation of *SSL* via MCMC or Variational inference similar to methods of (Hof and Speed, 2025), to enable a dense regression approach like VIPRS, which demonstrated strong predictive performance. Overall, we believe that *SSLPRS* provides effective analytical tools for predicting and understanding the genetic underpinnings of complex traits and diseases.

## Data Availability

All data produced in the present work are contained in the manuscript.

https://github.com/li-lab-mcgill/penprs

## Acknowledgments

We thank members of Li lab for their feedback and comments on earlier iterations of this work. Y.L. is supported by Canada Research Chair (Tier 2) in Machine Learning for Genomics and Healthcare (CRC-2021-00547) and Natural Sciences and Engineering Research Council (NSERC) Discovery Grant (RGPIN-2016-05174). This research used the NeuroHub infrastructure and was undertaken thanks in part to funding from the Canada First Research Excellence Fund, awarded through the Healthy Brains, Healthy Lives initiative at McGill University. This research was enabled in part by support provided by Calcul Québec and the Digital Research Alliance of Canada. This research has been conducted using the UK Biobank Resource under Application Number 45551. No competing interest declared.

