## Supplemental information for "Sparse Polygenic Risk Score Inference with the Spike-and-Slab LASSO"

### S1 Supplementary Figures

#### S1.1 Model variable selection performance across the warm-start ladder

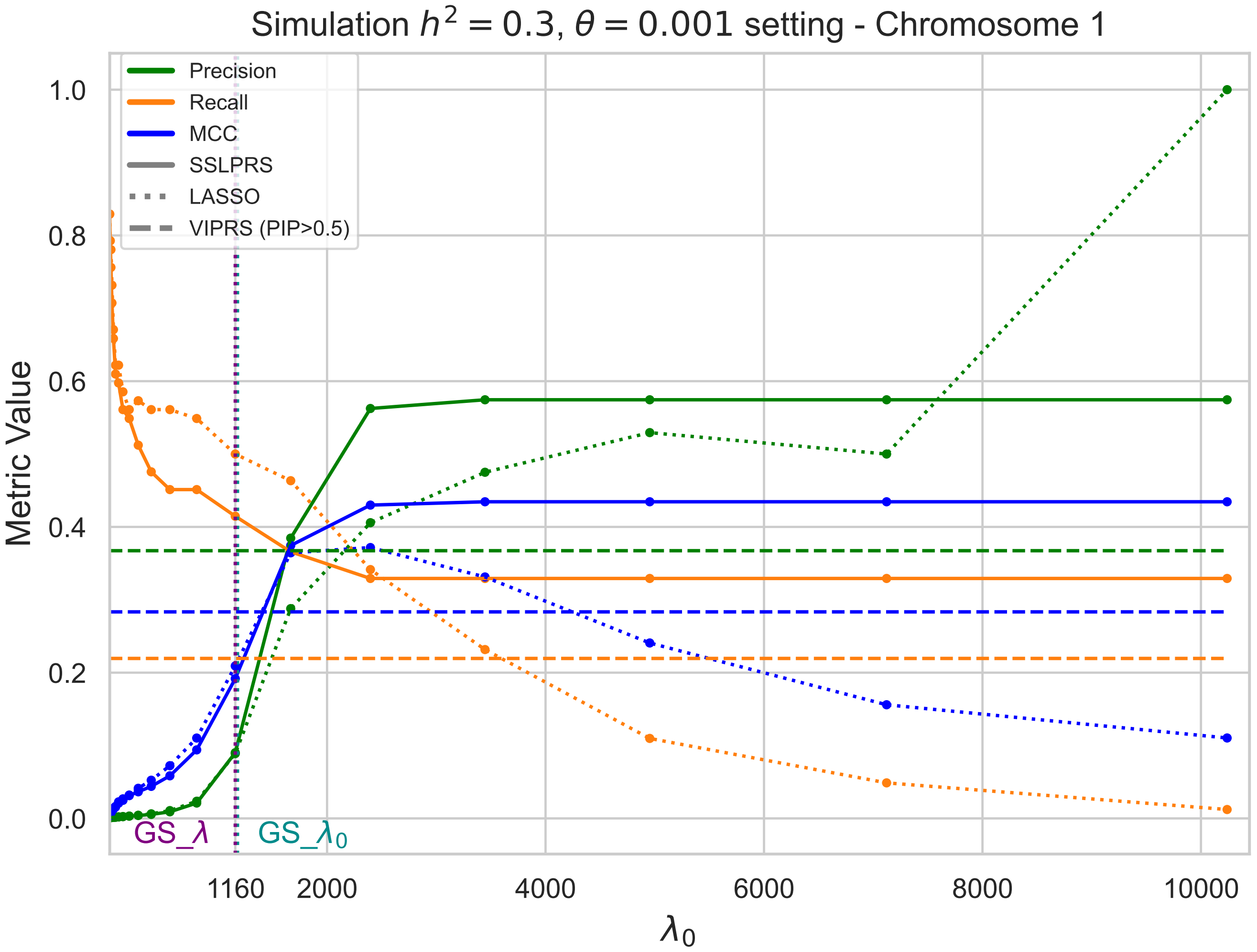


Figure S1: Variable selection performance across the $\lambda_0$ ladder under simulation setting of $h^2 = 0.3, \theta = 0.001$ on chromosome 1. Three variable selection metrics are shown in different colors: Precision (green), Recall (orange), Matthews Correlation Coefficient (MCC) (blue). The *LASSO* trends are shown as dotted lines, whereas the *SSLPRS* trends are shown as solid lines. For reference, the performance of the *VIPRS* model on these same metrics are shown as horizontal dashed lines. The 20 ladder steps of are indicated as scatter points. The vertical dotted cyan or purple line shows the grid-search-selected $\lambda_0$ or $\lambda$ by *SSLPRS* or *LASSO* respectively, annotated as "*GS_*$\lambda$" or "*GS_*$\lambda_0$". *VIPRS* uses the median rule, where variants with Posterior Inclusion Probability (PIP) > 0.5 are considered selected.

#### S1.2 Model Effect Size estimation against true effects in UK Biobank simulation

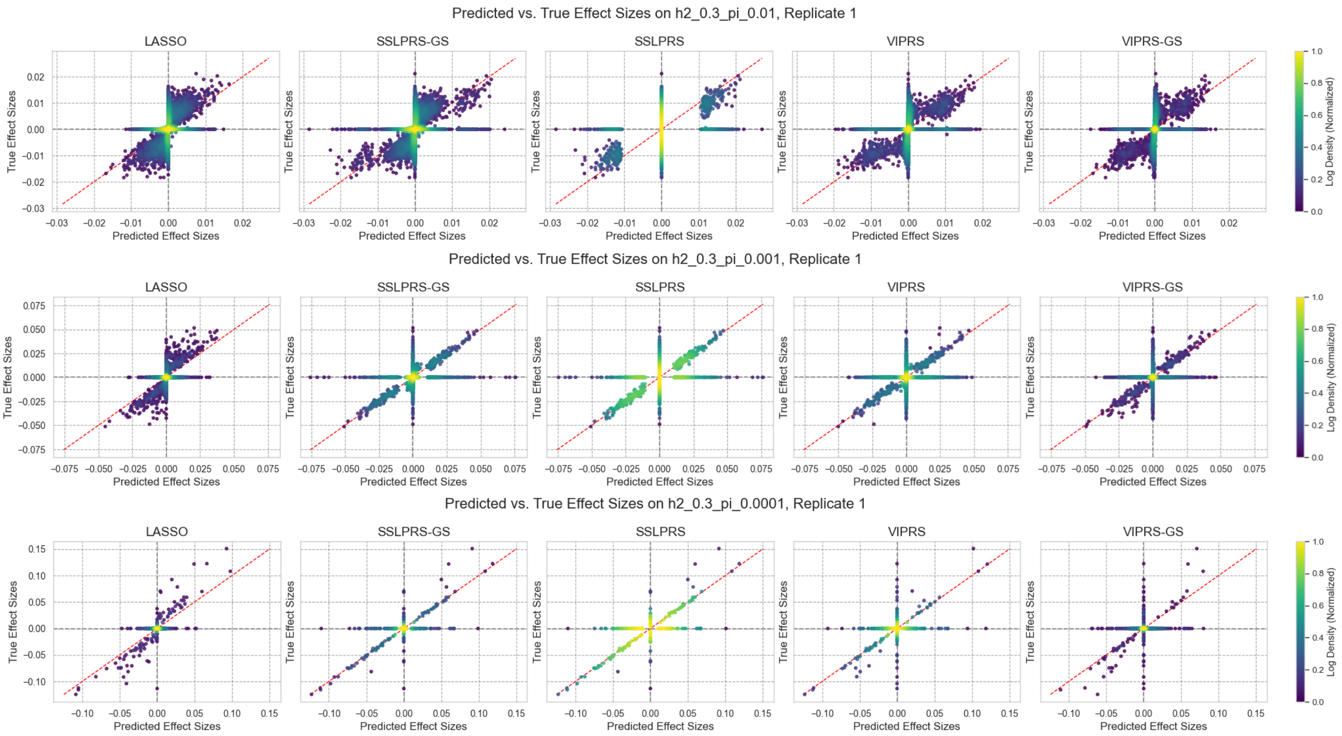


Figure S2: Model Effect Size estimation against true effects in UK Biobank simulation. Scatter plot of variants' predicted effect size (x-axis) estimates compared to true effect sizes (y-axis) on different sparse settings for causal proportion $\theta = \{0.01, 0.001, 0.0001\}$ on heritability $h^2 = 0.3$ of replicate 1. Scatter points are colored to show normalized log density. Variants with both predicted and true effects $< 5 \times 10^{-4}$ were excluded to reduce the influence of null associations and improve visualization of effect size densities of true or selected causal effect sizes. For reference, vertical $x = 0$ and horizontal $y = 0$ dotted lines shown in grey, y=x dotted line shown in red. The models shown from left to right are: *LASSO/SSLPRS-GS* (estimates of the best performing $\lambda/\lambda_0$ penalty on a 20-step pathwise/warm-start ladder), *SSLPRS* (estimates from fitting to the end of the 20-step warm-start ladder), *VIPRS/VIPRS-GS* (estimates from the EM inference version/best performing causal proportion $\theta$ across a ladder of 20 values.

#### S1.3 Pearson correlation on effect sizes of against true effects on UK Biobank simulation

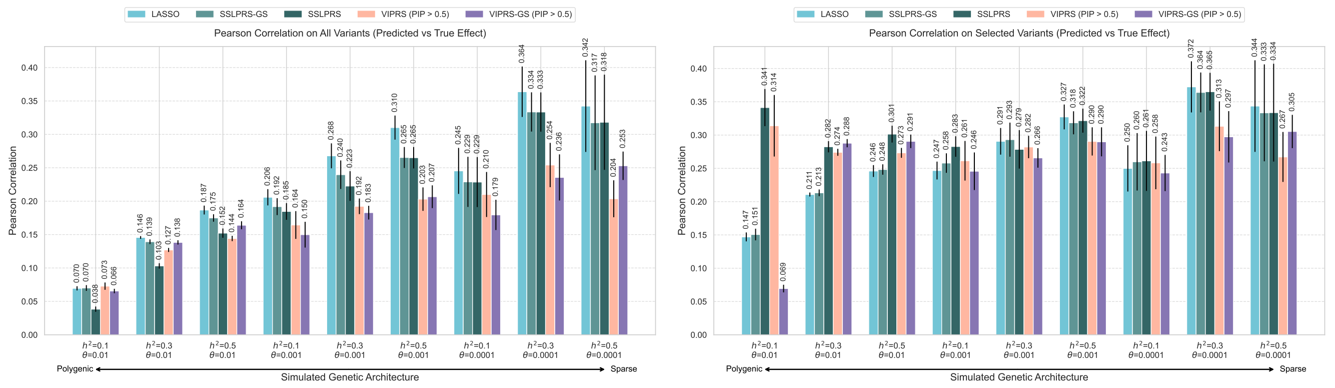


(a) (b)

Figure S3: Bar plot of the Pearson correlation between predicted and true effect sizes on (a) all variants, (b) selected variants, across simulated genomic architectures from polygenic causal proportion $\theta = 0.01$ to sparse settings of $\theta$=0.001, 0.0001 across different heritability $h^2$ values 0.1, 0.3, 0.5. For each architecture, bars for *LASSO, SSLPRS-GS, SSLPRS, VIPRS*, and *VIPRS-GS* are shown in different colors, displaying the average Pearson correlation across 5 replicates of the simulation setting. Vertical black bars show the standard error across the replicates. For (b), non-zero effect size estimates for the penalized models, *LASSO, SSLPRS, SSLPRS-GS* are considered as selected. As *VIPRS* do not produce exact sparse estimates, the median rule is used for selecting variants, where variants with a posterior inclusion probability (PIP) > 0.5 are considered selected.

#### S1.4 Coefficient paths of models across a ladder of penalty values on simulated UK Biobank Data.

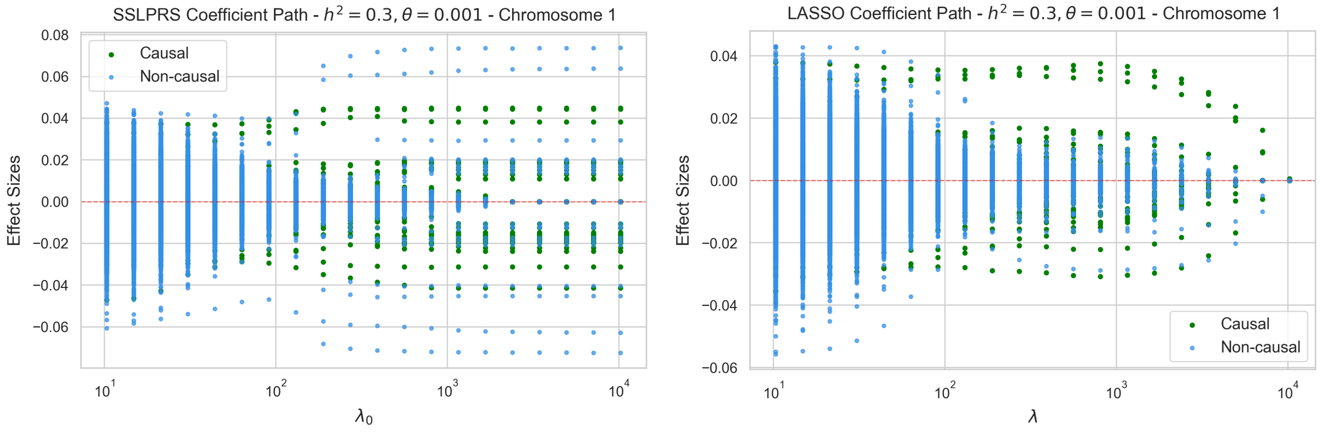


(a) (b)

Figure S4: Coefficient (Effect size) estimation path across the ladder of (a) $\lambda_0$ penalty values for the SSLPRS model, (b) $\lambda$ penalty values for the LASSO model on chromosome one of the balanced simulations setting with heritability $h^2 = 0.3$ and causal proportion $\theta = 0.001$. True non-causal variants are colored as light-blue, where causal variants are colored as green. Horizontal $y$ (effect size)= 0 dashed red line shown as reference. Penalty values across the ladder are shown on a log scale. (a) Effects are held steady by the slab component with an increasing number of causal variants going across the ladder and most negligible, non-causal variants are shrunk to 0 (b) All effect sizes are eventually shrunk to zero for LASSO.

#### S1.5 Model variable selection and predictive performance on mixture of normals simulations of UK Biobank Data

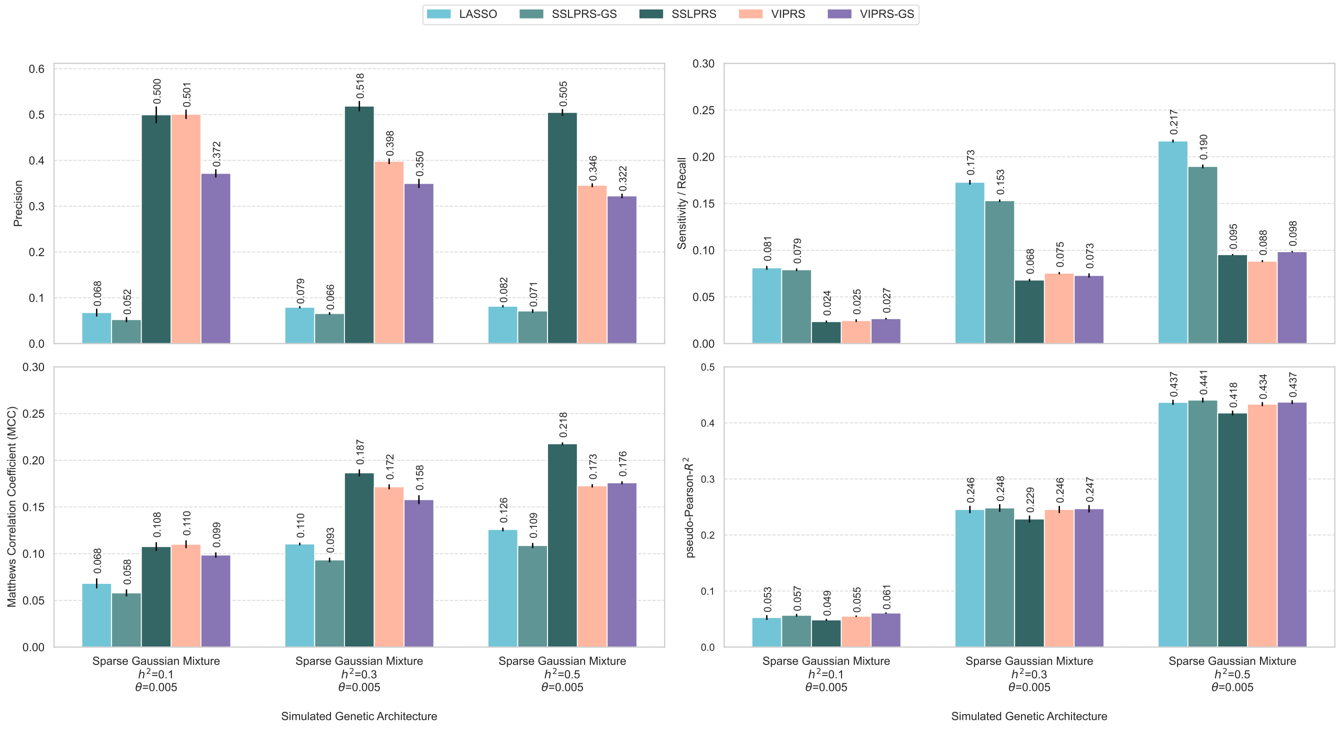


Figure S5: Variable selection and predictive performance of summary statistics-based PRS methods on simulated UK Biobank phenotypes through Mixture of Normals simulations. Performance is evaluated using four metrics: Precision (top-left), Recall/sensitivity (top-right), Matthews correlation coefficient (MCC, bottom-left), and Pearson-pseudo-$R^2$ (prediction accuracy, bottom-right) on held-out test set. The simulation configurations span three heritability settings, $h^2 = \{0.1, 0.3, 0.5\}$ with effects drawn from a Gaussian mixture prior with four components: Mixing proportions of $[0.995, 0.002, 0.002, 0.001]$, paired with variance multipliers $[0, 0.01, 0.1, 1]$. Under this specification, the causal proportion is $\theta = 0.005$ or 5%, with causal effects distributed across the three non-zero Gaussian components of increasing variance. Five summary statistics-based PRS methods are included in this figure: *LASSO*, our proposed *SSLPRS* and *SSLPRS-GS* (grid search), as well as *VIPRS* and *VIPRS-GS* (grid search). Bars represent the mean value over 5 independent replicates. The black vertical lines denote the standard error across the replicates. For the variable selection metrics, the median rule was applied for the *VIPRS* models, where variants with Posterior Inclusion Probability (PIP) $> 0.5$ were considered as selected.

#### S1.6 Coefficient paths and effect size scatter of LASSO and SSL on chromosome 5 of standing height data from UK Biobank.

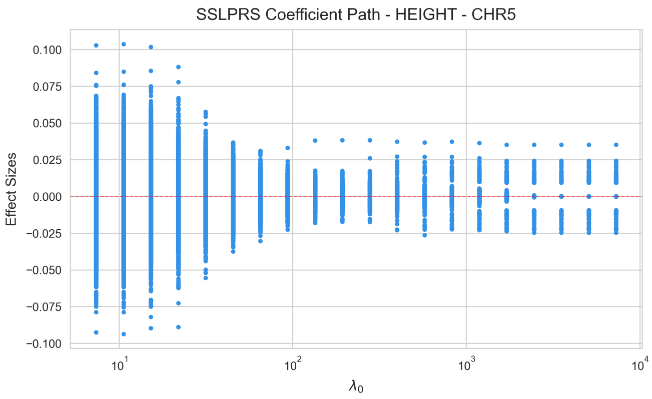


(a)

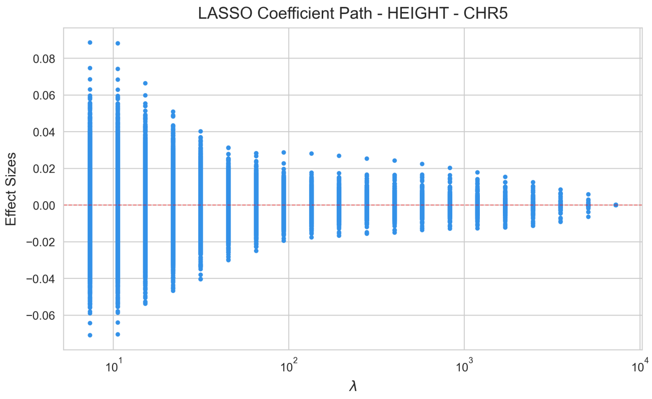


(b)

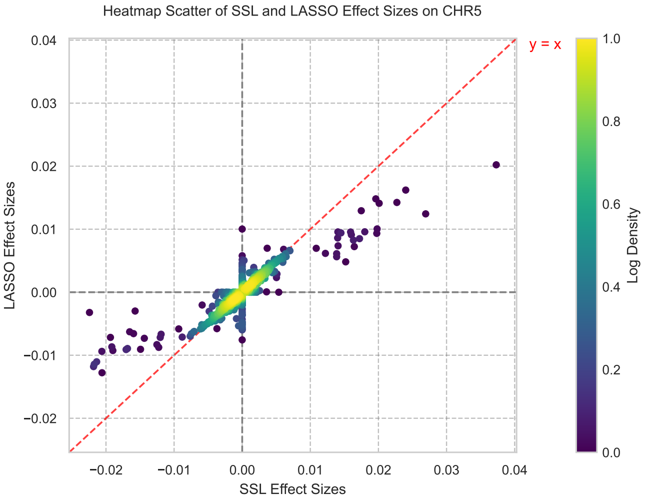


(c)

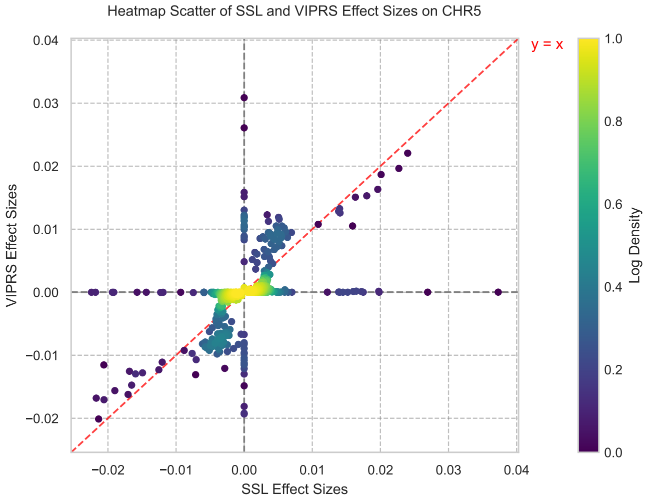


(d)

Figure S6: Coefficient paths and effect size scatter of *LASSO* and SSL on chromosome 5 of real height data from UK Biobank. (a,b) Coefficient (Effect size) estimation path across the ladder of $\lambda_0$, $\lambda$ penalty values for the *SSLPRS*, *LASSO* models respectively on chromosome 5 of the standing height (HEIGHT) data from the UK Biobank. Horizontal $y(\text{effect size}) = 0$ dashed red line shown as reference. (a) Variants thought to be causal according to the *SSLPRS* are held steady as variants assigned to be negligible by the model are gradually shrunk to zero. (b) Without a stabilizing slab, *LASSO* gradually shrinks all variants towards zero as the penalty increases along the ladder. (c, d) scatter plot of effect sizes of *SSLPRS* from the $\lambda_0$ value selected during grid-search against (c) *LASSO* effect size estimates from the lambda value selected during grid-search (d) *VIPRS* effect size estimates from EM inference. Variants with both predicted and true effects $< 5 \times 10^{-4}$ were excluded to reduce the influence of null associations and improve visualization of effect size densities of true or selected causal effect sizes. For reference, vertical $x = 0$ and horizontal $y = 0$ dotted lines shown in grey, $y = x$ dotted line shown in red.

#### S1.7 Effects of estimating variance on variable selection and prediction performance on UK Biobank simulation for *SSLPRS*

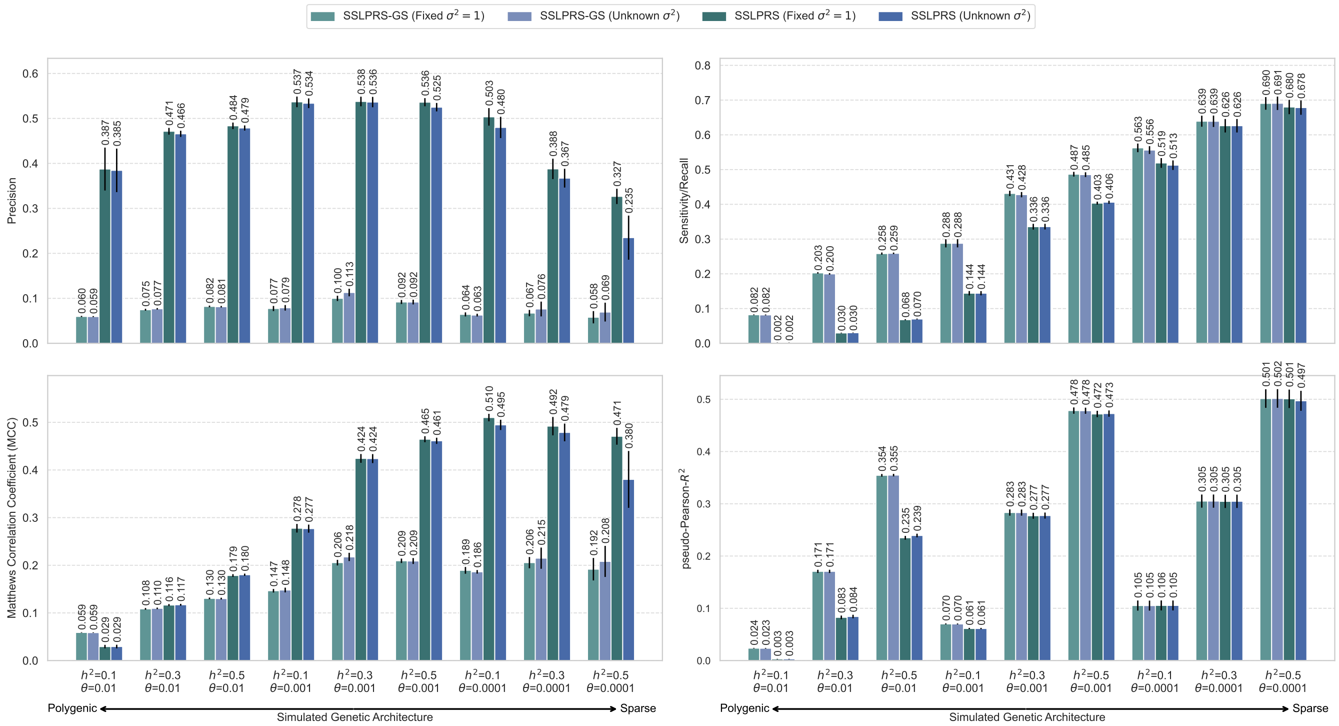


Figure S7: Variable selection and predictive performance of fixing and estimating variance for *SSLPRS* on simulated UK Biobank phenotypes. Performance is evaluated using four metrics: Precision (top-left), Recall/sensitivity (top-right), Matthews correlation coefficient (MCC, bottom-left), and pseudo-$R^2$ (prediction accuracy, bottom-right) on held-out test set. The 9 simulation configurations span three heritability settings, $h^2 = \{0.1, 0.3, 0.5\}$, and three proportions of causal variants, $\theta = \{0.01, 0.001, 0.0001\}$. The proposed *SSLPRS* and *SSLPRS-GS* models are shown to compare the effects of fixing variance $\sigma^2 = 1$, or estimating it along the warm-start $\lambda_0$ spike penalty ladder. Bars represent the mean value over 5 independent replicates. The black vertical lines denote the standard error across the replicates.

#### S1.8 Alpha heritability modeled predictive performance on real quantitative phenotypes in the UK Biobank

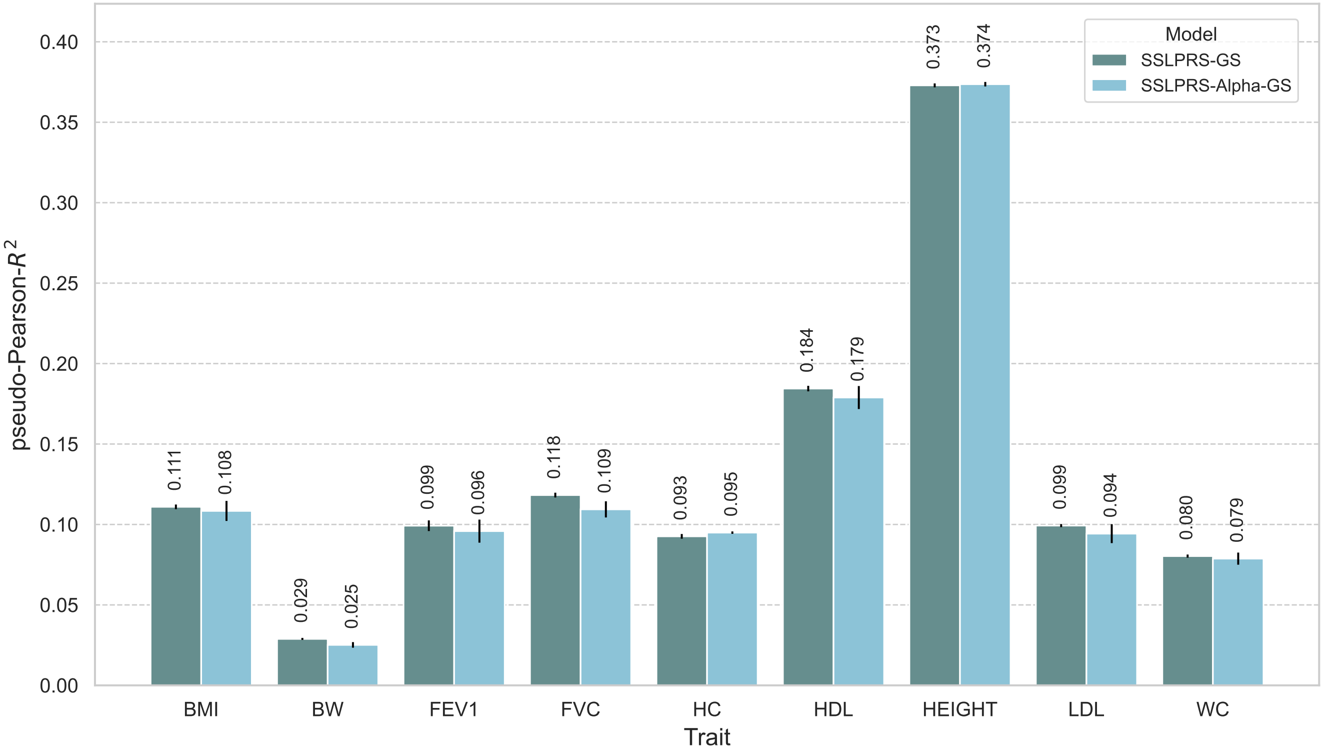


Figure S8: Predictive performance of grid searched *SSLPRS-GS* and its alpha-heritability modeled variant *SSLPRS-Alpha-GS*, with $\alpha = -0.25$, on real quantitative phenotypes in the UK Biobank. Prediction accuracy (Pseudo-Pearson-$R^2$) on held-out test sets in an analysis of 9 quantitative phenotypes: standing height (HEIGHT), high-density lipoprotein (HDL), body mass index (BMI), forced vital capacity (FVC), forced expiratory volume in 1 s (FEV1), hip circumference (HC), waist circumference (WC), low-density lipoprotein (LDL), and birth weight (BW). The bars represent the average model performance based on 5-fold cross-validation, with black vertical lines indicating the associated standard errors.

#### S1.9 Model predictive performance of real quantitative phenotypes with *Bersia* `LDetect` blocks and *SBayesRC* merged LD blocks

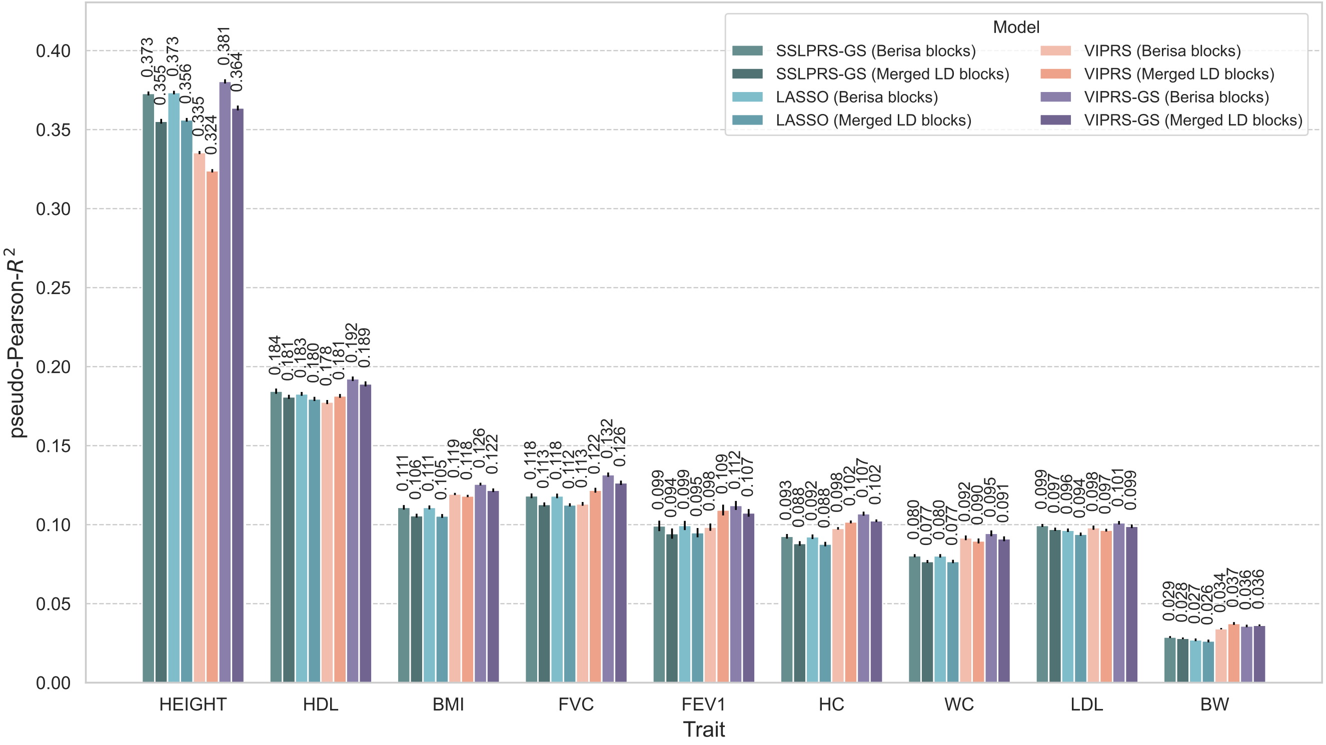


Figure S9: Predictive performance of summary statistics-based PRS methods on real quantitative phenotypes in the UK Biobank using two LD block schemes: *Bersia* `LDetect` blocks used by LD reference for the models in the analyses and the *SBayesRC* merged LD blocks. Prediction accuracy (Pseudo-Pearson-$R^2$) on held-out test sets in an analysis of 9 quantitative phenotypes: standing height (HEIGHT), high-density lipoprotein (HDL), body mass index (BMI), forced vital capacity (FVC), forced expiratory volume in 1 s (FEV1), hip circumference (HC), waist circumference (WC), low-density lipoprotein (LDL), and birth weight (BW). The bars represent the average model performance based on 5-fold cross-validation, with black vertical lines indicating the associated standard errors.

#### S1.10 Model predictive performance of real quantitative phenotypes in minority populations from the UK Biobank

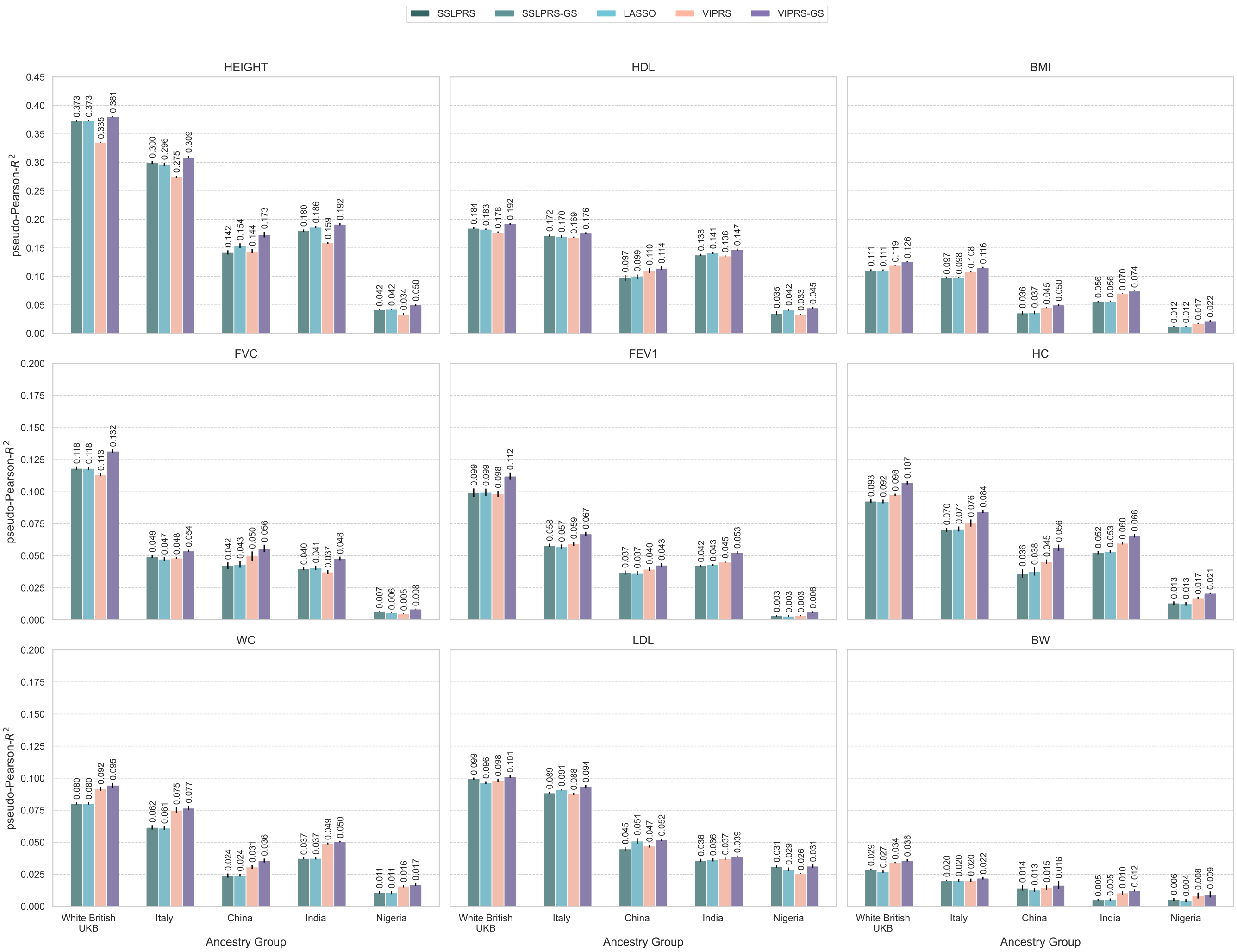


Figure S10: Predictive performance of summary statistics-based PRS methods on real quantitative phenotypes in minority populations from the UK Biobank. Effect size estimates were obtained from model training on summary statistics of the White British cohort and subsequently transferred to minority ancestry groups and evaluated on the summary statistics minority of ancestry groups of Italy, China, India, and Nigeria. Prediction accuracy (Pseudo-Pearson-$R^2$) on held-out test sets in an analysis of 9 quantitative phenotypes: standing height (HEIGHT), high-density lipoprotein (HDL), body mass index (BMI), forced vital capacity (FVC), forced expiratory volume in 1 s (FEV1), hip circumference (HC), waist circumference (WC), low-density lipoprotein (LDL), and birth weight (BW) showed per panel. The bars represent the average model performance based on 5-fold cross-validation, with black vertical lines indicating the associated standard errors.

#### S1.11 Effects of $\lambda_{min}$ on model training stability and prediction performance

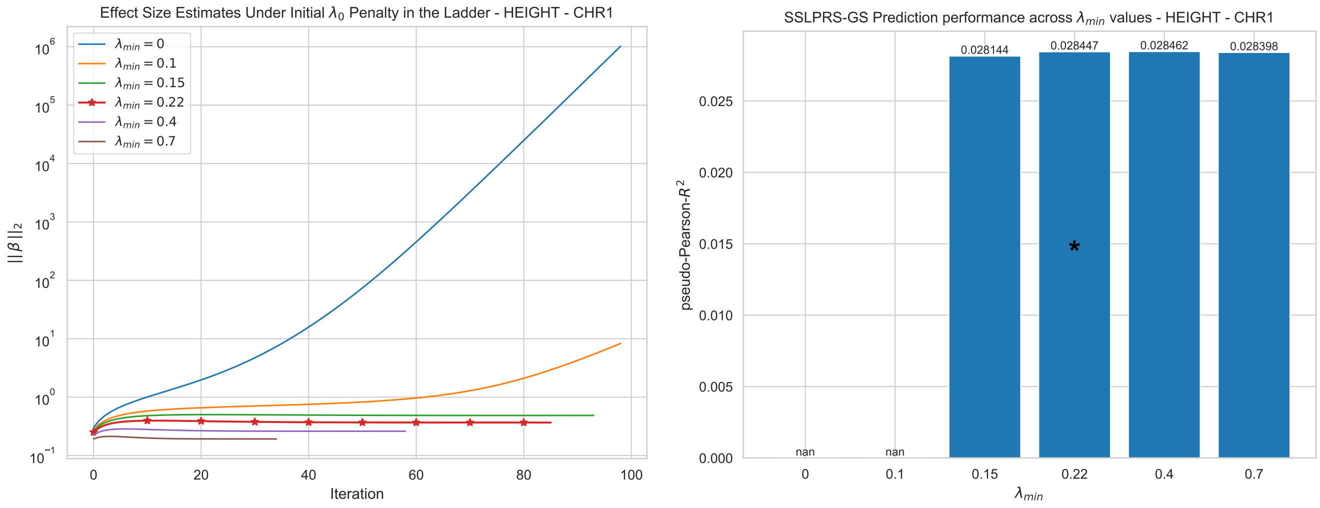


(a) (b)

Figure S11: Effects of $\lambda_{\mathrm{min}}$ on training stability and prediction performance for chromosome 1 of standing height from the UK Biobank. `int8` quantized LD matrices are used to better illustrate the difference between $\lambda_{min}$ values. (a) Stability of effect size estimates, measured by $||\beta||_2$, on the initial spike $\lambda_0$ penalty of the warm-start $\lambda_0$ ladder across different $\lambda_{min}$ values, shown in different colors. The provided $\lambda_{min} = 0.22$ value based on LD matrix properties, provided through the `magenpy` package is starred. (b) Predictive performance shown in pseudo-Pearson-$R^2$ of the best performing *SSLPRS* model grid searched on the warm-start ladder of $\lambda_0$ spike penalties (*SSLPRS-GS*). Models with diverging or unstable training are shown as "nan". The bar of the provided $\lambda_{min} = 0.22$ value based on LD matrix properties, provided through the `magenpy` package is starred.

#### S1.12 Effect size estimates comparison between *LASSO* and *Lassosum* across different $\lambda$ penalty and LD scaling hyperparameter $s$

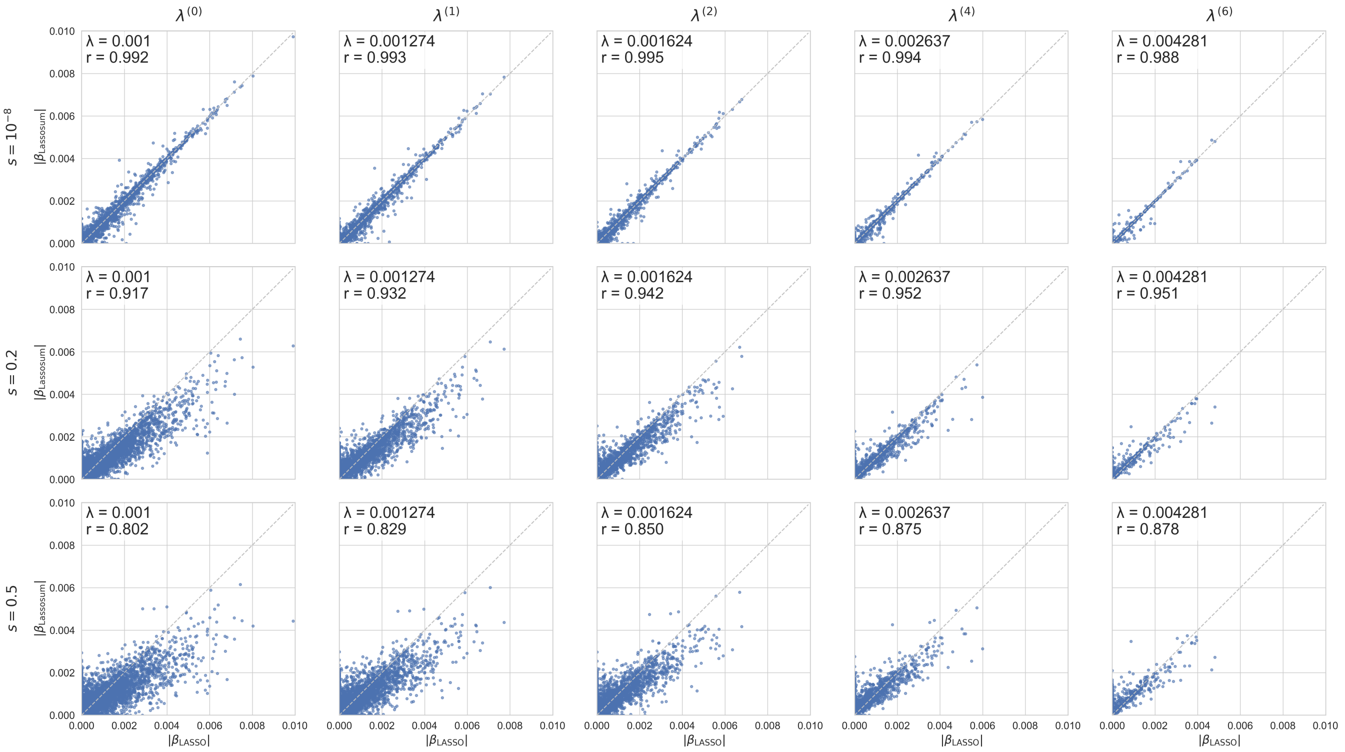


Figure S12: Effect size estimates comparison between *LASSO* and *Lassosum* across different $\lambda$ penalty and LD scaling hyperparameter $s$ on chromosome 22 of Body Mass Index (BMI) phenotype from the UK Biobank. Grid of scatter plots shown across $\lambda$ penalty indices $[0, 1, 2, 4, 6]$ of the default 20-step log ladder from 0.001 to 0.01 as used in *Lassosum* and different LD scaling hyperparameter $s = [10^{-8}, 0.2, 0.5]$ used in *Lassosum*. *LASSO* penalties were standardized to correspond with the penalty ladder of *Lassosum*. Scatter plots display the absolute value of effect size estimates of *LASSO* (x-axis) against the estimates of *Lassosum* (y-axis). The y=x line is show as a dotted gray line. The $\lambda$ penalty magnitude per index along with the correlation of the effect size estimates for the two models are annotated per scatter.

#### S1.13 Regression calibration of predicted PRS across quantiles on UK Biobank standing height

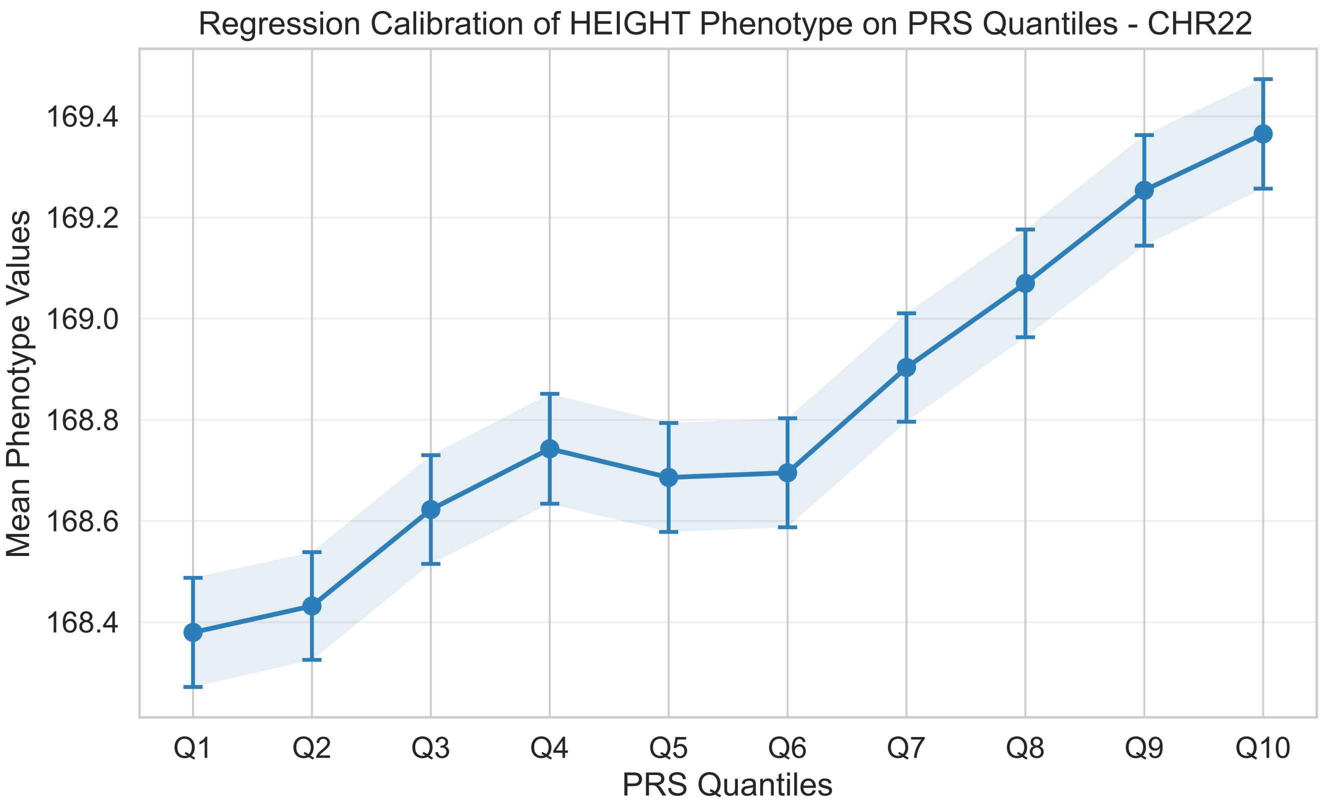


Figure S13: Regression calibration of standing height phenotype from UK Biobank on chromosome 22 for *SSLPRS-GS*. Individuals were ranked by predicted polygenic risk scores (PRS) and stratified into ten equal-sized predicted PRS quantiles (Q1 low - Q10 high) for mean phenotype (height) value OLS regression per quantile. Points indicate the regressed mean phenotypes. The standard error bar is shown with a light blue shaded region across the quantiles.

#### S1.14 Computational performance of summary statistics-based PRS methods on real quantitative phenotypes in the UK Biobank with up to 1 million genetic variants

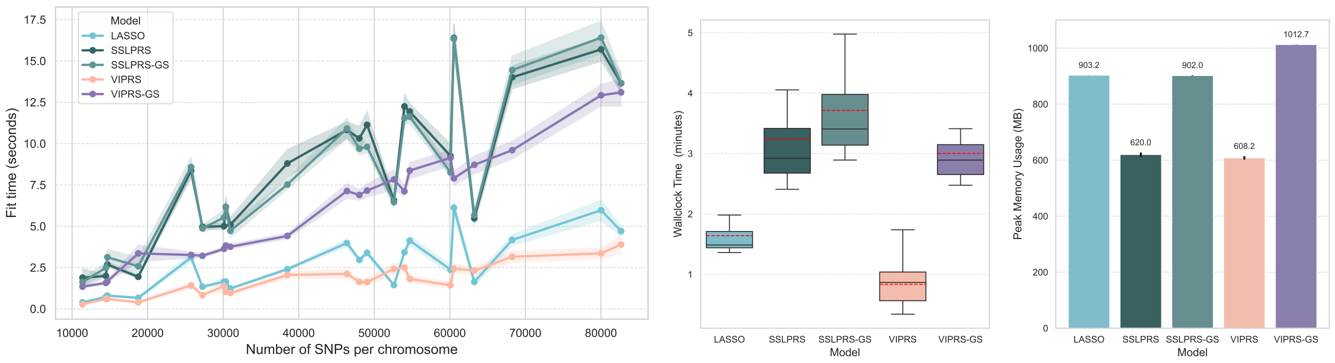


(a) (b) (c)

Figure S14: Computational performance of summary statistics-based PRS methods on real quantitative phenotypes in the UK Biobank with up to 1 million genetic variants. All metrics are shown as aggregate statistics across 5 folds and 9 quantitative real phenotypes from the UK Biobank, for a total of 45 experiments. Models are differentiated by color. For simplicity, *SSLPRS* and *SSLPRS-GS* are grouped together, as both require fitting to the end of the $\lambda_0$ ladder and incur similar computational costs. Panel (a) shows inference time per chromosome (in seconds). Chromosomes are represented on the x-axis in terms of the number of variants, scatter points show the average runtime and shaded regions show interpolated standard errors. (b) Boxplot of total Wallclock time (in minutes) for each of the four models. The thick horizontal block line shows the median and the red dotted line shows the mean wallclock time for each model. (c) Average Peak Memory utilization (in Megabytes) for each of the four models. Small vertical block lines on top of the bars show standard errors

### S2 Supplementary Tables

#### S2.1 Variable selection performance of *SSLPRS* on simulated phenotypes with different genetic architectures.

| Setting | SSLPRS-GS | | | SSLPRS | | |
|---|---|---|---|---|---|---|
| | Precision | Sensitivity | MCC | Precision | Sensitivity | MCC |
| $h^2 = 0.1$<br>$\theta = 0.01$ | $0.0597 \pm 0.0014$ | $\mathbf{0.0822 \pm 0.0012}$ | $\mathbf{0.0590 \pm 0.0006}$ | $\mathbf{0.3874 \pm 0.0533}$ | $0.0023 \pm 0.0003$ | $0.0293 \pm 0.0041$ |
| $h^2 = 0.3$<br>$\theta = 0.01$ | $0.0747 \pm 0.0023$ | $\mathbf{0.2028 \pm 0.0020}$ | $0.1085 \pm 0.0020$ | $\mathbf{0.4701 \pm 0.0086}$ | $0.0297 \pm 0.0009$ | $\mathbf{0.1162 \pm 0.0023}$ |
| $h^2 = 0.5$<br>$\theta = 0.01$ | $0.0820 \pm 0.0021$ | $\mathbf{0.2584 \pm 0.0036}$ | $0.1305 \pm 0.0020$ | $\mathbf{0.4837 \pm 0.0082}$ | $0.0680 \pm 0.0015$ | $\mathbf{0.1786 \pm 0.0033}$ |
| $h^2 = 0.1$<br>$\theta = 0.001$ | $0.0774 \pm 0.0063$ | $\mathbf{0.2880 \pm 0.0132}$ | $0.1466 \pm 0.0041$ | $\mathbf{0.5388 \pm 0.0132}$ | $0.1446 \pm 0.0069$ | $\mathbf{0.2786 \pm 0.0099}$ |
| $h^2 = 0.3$<br>$\theta = 0.001$ | $0.1000 \pm 0.0066$ | $\mathbf{0.4316 \pm 0.0085}$ | $0.2056 \pm 0.0066$ | $\mathbf{0.5405 \pm 0.0133}$ | $0.3356 \pm 0.0093$ | $\mathbf{0.4254 \pm 0.0109}$ |
| $h^2 = 0.5$<br>$\theta = 0.001$ | $0.0889 \pm 0.0030$ | $\mathbf{0.4862 \pm 0.0076}$ | $0.2061 \pm 0.0025$ | $\mathbf{0.5361 \pm 0.0086}$ | $0.4047 \pm 0.0043$ | $\mathbf{0.4653 \pm 0.0061}$ |
| $h^2 = 0.1$<br>$\theta = 0.0001$ | $0.0642 \pm 0.0050$ | $\mathbf{0.5625 \pm 0.0139}$ | $0.1892 \pm 0.0079$ | $\mathbf{0.5025 \pm 0.0218}$ | $0.5190 \pm 0.0156$ | $\mathbf{0.5095 \pm 0.0084}$ |
| $h^2 = 0.3$<br>$\theta = 0.0001$ | $0.0671 \pm 0.0079$ | $\mathbf{0.6392 \pm 0.0177}$ | $0.2054 \pm 0.0132$ | $\mathbf{0.3883 \pm 0.0254}$ | $0.6262 \pm 0.0217$ | $\mathbf{0.4923 \pm 0.0217}$ |
| $h^2 = 0.5$<br>$\theta = 0.0001$ | $0.0580 \pm 0.0152$ | $\mathbf{0.6905 \pm 0.0203}$ | $0.1917 \pm 0.0263$ | $\mathbf{0.3261 \pm 0.0190}$ | $0.6801 \pm 0.0230$ | $\mathbf{0.4703 \pm 0.0197}$ |

Table S1: Detailed variable selection performance comparison of our proposed *SSLPRS* models on simulated phenotypes of UK Biobank data with different genetic architectures. The models shown are: the default *SSLPRS* and *SSLPRS-GS*. The 9 simulation configurations span three heritability settings, $h^2 = \{0.1, 0.3, 0.5\}$, and three proportions of causal variants, $\theta = \{0.01, 0.001, 0.0001\}$. The metrics shown are precision, recall, and the Matthews Correlation Coefficient(MCC). The model performance of these metrics are reported as the mean $\pm$ the standard error across 5 different replicates.

#### S2.2 Hyperparameter ablation analysis of *SSLPRS* and *SSLPRS-GS*

| Model | Hyperparameter | Value | Pearson-pseudo-$R^2$ | Precision | Recall | MCC |
|---|---|---|---|---|---|---|
| **Effects of Prior Mean on Mixing Proportion $\theta$ (tuned through $a$)** | | | | | | |
| SSLPRS | $\mathbb{E}(\theta)$ | 0.01 | 0.270 | **0.553** | 0.316 | 0.418 |
| SSLPRS | $\mathbb{E}(\theta)$ | <u>0.05</u> | 0.277 | 0.542 | 0.343 | **0.431** |
| SSLPRS | $\mathbb{E}(\theta)$ | 0.10 | 0.277 | 0.523 | 0.342 | 0.422 |
| SSLPRS | $\mathbb{E}(\theta)$ | 0.15 | 0.278 | 0.509 | 0.343 | 0.417 |
| SSLPRS | $\mathbb{E}(\theta)$ | 0.20 | **0.279** | 0.501 | **0.347** | 0.417 |
| SSLPRS-GS | $\mathbb{E}(\theta)$ | 0.01 | 0.282 | 0.097 | **0.434** | 0.204 |
| SSLPRS-GS | $\mathbb{E}(\theta)$ | <u>0.05</u> | **0.284** | 0.101 | 0.423 | 0.205 |
| SSLPRS-GS | $\mathbb{E}(\theta)$ | 0.10 | **0.284** | 0.119 | 0.410 | 0.219 |
| SSLPRS-GS | $\mathbb{E}(\theta)$ | 0.15 | 0.283 | 0.124 | 0.404 | 0.222 |
| SSLPRS-GS | $\mathbb{E}(\theta)$ | 0.20 | 0.283 | **0.140** | 0.398 | **0.235** |
| **Effects of scale hyperparameter $\epsilon$ for $\lambda_1$, ($\lambda_1 = \epsilon\lambda_{\max}$)** | | | | | | |
| SSLPRS | $\epsilon$ | 0.0001 | 0.265 | 0.571 | 0.311 | 0.421 |
| SSLPRS | $\epsilon$ | <u>0.001</u> | 0.277 | 0.542 | 0.343 | **0.431** |
| SSLPRS | $\epsilon$ | 0.010 | **0.280** | 0.493 | **0.348** | 0.414 |
| SSLPRS | $\epsilon$ | 0.100 | 0.271 | 0.547 | 0.322 | 0.420 |
| SSLPRS | $\epsilon$ | 0.250 | 0.223 | **0.606** | 0.219 | 0.364 |
| SSLPRS-GS | $\epsilon$ | 0.0001 | 0.280 | 0.083 | **0.457** | 0.193 |
| SSLPRS-GS | $\epsilon$ | <u>0.001</u> | **0.284** | 0.101 | 0.423 | 0.205 |
| SSLPRS-GS | $\epsilon$ | 0.010 | **0.284** | 0.154 | 0.406 | 0.249 |
| SSLPRS-GS | $\epsilon$ | 0.100 | 0.281 | 0.095 | 0.444 | 0.204 |
| SSLPRS-GS | $\epsilon$ | 0.250 | 0.256 | **0.295** | 0.411 | **0.348** |
| **Effects of $\lambda_0$ ladder length** | | | | | | |
| SSLPRS | Steps | 10 | 0.276 | 0.527 | 0.337 | 0.421 |
| SSLPRS | Steps | <u>20</u> | **0.277** | **0.542** | 0.343 | **0.431** |
| SSLPRS | Steps | 30 | 0.275 | **0.542** | 0.337 | 0.427 |
| SSLPRS-GS | Steps | 10 | 0.279 | **0.249** | 0.368 | **0.302** |
| SSLPRS-GS | Steps | <u>20</u> | **0.284** | 0.101 | 0.423 | 0.205 |
| SSLPRS-GS | Steps | 30 | 0.283 | 0.097 | **0.428** | 0.202 |

Table S2: Ablation study of our proposed *SSLPRS* and *SSLPRS-GS* under variations of key hyperparameters on a replicate of the balanced simulated setting: heritability $h^2 = 0.3$, causal proportion $\theta = 0.001$. We conducted three sets of ablations: (i) the effects of the prior mean $\mathbb{E}(\theta) = [0.01, 0.05, 0.10, 0.15, 0.2]$ on the mixing proportion $\theta$, tuned through the hyperparameter $a$; (ii) the effects of the hyperparameter $\epsilon = [0.0001, 0.001, 0.01, 0.10, 0.25]$, which scales the slab penalty $\lambda_1$ with respect to $\lambda_{max}$; (iii) the effects of different spike penalty $\lambda_0$ ladder length $= [10, 20, 30]$. The predictive accuracy is shown in pseudo-Pearson-$R^2$, alongside variable selection performance in precision, recall, and Matthews Correlation Coefficient(MCC). The default model setting is underlined and the best-performing values per model are highlighted in bold.

#### S2.3 Model replication performance of selected variants in the GLGC consortium meta-analysis

| Trait | Model | # of GLGC Significant Variants | # of Variants Selected by Model | # of Replicated Variants | Significance MCC | Replication rate |
|---|---|---|---|---|---|---|
| LDL | LASSO | 8479 | $10079 \pm 437$ | $884 \pm 9$ | $0.087 \pm 0.001$ | $0.088 \pm 0.003$ |
| LDL | SSLPRS-GS | 8479 | $8453 \pm 196$ | $615 \pm 7$ | $0.065 \pm 0.002$ | $0.073 \pm 0.002$ |
| LDL | SSLPRS | 8479 | $180 \pm 3$ | $157 \pm 3$ | $0.126 \pm 0.001$ | $\mathbf{0.871 \pm 0.005}$ |
| LDL | VIPRS | 8479 | $249 \pm 5$ | $194 \pm 4$ | $\mathbf{0.133 \pm 0.002}$ | $0.782 \pm 0.008$ |
| LDL | VIPRS-GS | 8479 | $61720 \pm 16627$ | $539 \pm 124$ | $0.025 \pm 0.028$ | $0.156 \pm 0.148$ |
| HDL | LASSO | 6677 | $23775 \pm 707$ | $1101 \pm 6$ | $0.076 \pm 0.002$ | $0.046 \pm 0.002$ |
| HDL | SSLPRS-GS | 6677 | $22933 \pm 929$ | $710 \pm 10$ | $0.045 \pm 0.001$ | $0.031 \pm 0.001$ |
| HDL | SSLPRS | 6677 | $310 \pm 3$ | $226 \pm 3$ | $\mathbf{0.156 \pm 0.002}$ | $\mathbf{0.729 \pm 0.011}$ |
| HDL | VIPRS | 6677 | $582 \pm 4$ | $300 \pm 3$ | $0.150 \pm 0.001$ | $0.515 \pm 0.004$ |
| HDL | VIPRS-GS | 6677 | $73530 \pm 23296$ | $547 \pm 135$ | $0.005 \pm 0.006$ | $0.011 \pm 0.004$ |

Table S3: Model Replication Performance of Selected Variant in the Global Lipids Genetics Consortium (GLGC) Consortium Meta-analysis (Graham et al., 2021) for low- and high-density lipoprotein (HDL, LDL) phenotypes. Five summary statistics-based models are included: *LASSO*, our proposed SSLPRS and *SSLPRS-GS* (grid search), as well as *VIPRS* and *VIPRS-GS* (grid search). GLGC variants that have p-values $< 5 \times 10^{-8}$ are considered significant. For the penalized models (*LASSO, SSLPRS*), variants are considered selected its effect size is nonzero, and for Bayesian models like *VIPRS*, the median rule was applied for variable selection, retaining only variants with Posterior Inclusion Probability (PIP) $> 0.5$. Replicated variants are the selected variants by the model that is also considered significant in the GLGC meta-analysis. The significance Matthews Correlation Coefficient (MCC) is defined as $MCC = (TP \times TN - FP \times FN)/(\sqrt{(TP + FP)(TP + FN)(TN + FP)(TN + FN)})$, where $TP$ (true positives) denotes variants selected by the model and is also significant in GLGC, $TN$ (true negatives), $FP$ (false positives), and $FN$ (false negatives) are defined accordingly. Replication rate indicate the proportion of selected variants by the model that is also considered significant significant in GLGC, i.e. `# of Replicated Variants / # of Variants Selected by Model`. The average across the 5-folds is indicated followed by the standard error. Metrics pertaining to the number of variants are rounded to the nearest whole number. For each trait, the best performing model in terms of significance MCC and replication rate is shown in bold.

#### S2.4 Sensitivity of *VIPRS* models on PIP thresholds for variable selection and replication performance

| Model | PIP thr. | Selected Variants Count | FDR-controlled replication count | FDR-controlled replication rate |
|---|---|---|---|---|
| **VIPRS** | 0.3 | $276 \pm 6$ | $\mathbf{254 \pm 6}$ | $0.920 \pm 0.009$ |
| | <u>0.5</u> | $249 \pm 12$ | $235 \pm 10$ | $0.946 \pm 0.010$ |
| | 0.7 | $222 \pm 8$ | $217 \pm 7$ | $0.975 \pm 0.012$ |
| | 0.9 | $\mathbf{193 \pm 6}$ | $190 \pm 6$ | $\mathbf{0.989 \pm 0.007}$ |
| **VIPRS-GS** | 0.3 | 64,669 $\pm$ 37,018 | **1,729 $\pm$ 956** | $0.188 \pm 0.362$ |
| | <u>0.5</u> | 61,720 $\pm$ 37,180 | 1,658 $\pm$ 963 | $0.198 \pm 0.386$ |
| | 0.7 | 41,977 $\pm$ 38,471 | 1,344 $\pm$ 1,093 | $0.397 \pm 0.502$ |
| | 0.9 | **41,941 $\pm$ 38,473** | 1,313 $\pm$ 1,091 | $\mathbf{0.409 \pm 0.518}$ |

| Model | PIP thr. | Precision | Recall | MCC |
|---|---|---|---|---|
| **VIPRS** | 0.3 | $0.341 \pm 0.076$ | $\mathbf{0.252 \pm 0.176}$ | $\mathbf{0.255 \pm 0.113}$ |
| | <u>0.5</u> | $0.347 \pm 0.075$ | $0.248 \pm 0.177$ | $\mathbf{0.255 \pm 0.118}$ |
| | 0.7 | $\mathbf{0.348 \pm 0.073}$ | $0.242 \pm 0.177$ | $0.252 \pm 0.122$ |
| | 0.9 | $0.347 \pm 0.073$ | $0.234 \pm 0.178$ | $0.247 \pm 0.126$ |
| **VIPRS-GS** | 0.3 | $0.244 \pm 0.125$ | $\mathbf{0.292 \pm 0.156}$ | $0.213 \pm 0.098$ |
| | <u>0.5</u> | $0.261 \pm 0.131$ | $0.287 \pm 0.157$ | $\mathbf{0.218 \pm 0.102}$ |
| | 0.7 | $0.268 \pm 0.131$ | $0.277 \pm 0.156$ | $\mathbf{0.218 \pm 0.105}$ |
| | 0.9 | $\mathbf{0.277 \pm 0.132}$ | $0.269 \pm 0.161$ | $0.216 \pm 0.110$ |

Table S4: Sensitivity of Posterior Inclusion Probability (PIP) thresholds for variable selection and replication performance on the *VIPRS* models. PIP thresholds 0.3, 0.5, 0.7, 0.9 are shown. The replication metrics for models trained on UK Biobank data are shown in the top half of the table, where replication was assessed based on statistical p-value significance in the independent Global Lipids Genetics Consortium (GLGC) meta-analysis for the LDL trait: number of selected variants by the model using the respective PIP threshold, the Benjamini–Hochberg False Discovery Rate (FDR) $\alpha = 0.05$ controlled replication count (rounded to the nearest whole number) and replication rate. The variable selection metrics on the nine simulated UK Biobank phenotypes across genetic architecture combinations of causal proportion $\theta = [0.01, 0.001, 0.0001]$ and heritability $h^2 = [0.1, 0.3, 0.5]$ are shown in the lower half of the table: precision, recall, and the Matthew Correlation Coefficient (MCC). The mean and the standard deviation (mean $\pm$ sd) across the 5 cross validation folds, and across the 5 replicates are shown for the replication metrics and variable selection metrics respectively. The best performing PIP Threshold for each metric, per model, is bolded and the default choice of $PIP > 0.5$ (median rule) is underlined.

#### S2.5 Computational performance comparison of summary statistics based ***SSLPRS*** against individual level based `SSLASSO`

| $n$ | $p$ | *SSLPRS* | | `SSLASSO` individual | |
|---|---|---|---|---|---|
| | | Time (s) | PMU (MB) | Time (s) | PMU(MB) |
| 10000 | 5000 | 3.10 | 56.4 | 259.03 | 961.8 |
| 10000 | 10000 | 5.76 | 72.6 | 561.53 | 1934.3 |
| 10000 | 16296 | 10.11 | 89.9 | 864.93 | 3139.6 |
| 25000 | 5000 | 2.29 | 56.4 | 673.22 | 2389.3 |
| 25000 | 10000 | 3.88 | 72.6 | 1409.36 | 4777.2 |
| 25000 | 16296 | 7.48 | 89.8 | 2667.05 | 6236.3 |
| 50000 | 5000 | 1.85 | 56.4 | 1419.00 | 4763.8 |
| 50000 | 10000 | 3.92 | 72.6 | 3098.53 | 9530.0 |
| 50000 | 16296 | 5.00 | 89.9 | 4269.00 | 12503.7 |
| Average | – | 5.29 | 72.95 | 1343.2 | 4099.3 |

Table S5: Comparison of inference time (seconds) and peak memory usage (PMU, MB) between summary statistics-based *SSLPRS* and individual-level `SSLASSO` implementation in `R/Rcpp` (Ročková and George, 2018). We compare computational performance while varying sample sizes ($n$) in the UK Biobank and number of variants ($p$) on chromosome 22. For each sample size $n \in \{10000, 25000, 50000\}$, the number of SNPs is scaled from $p = 5000$ to $p = 10000$ and to the full chromosome $p = 16296$. The average inference time and peak memory usage of the two models are shown as well.

### S3 Supplementary Methods

#### S3.1 Data simulation and real data preparation

##### S3.1.1 Simple data simulation

The data for the simple simulation study, shown visually in Figure 1c of the main text, were simulated with a sample size of $n = 50$ on $p = 24$ SNPs. The genotype matrix follows a multivariate Gaussian $\boldsymbol{X} \sim \mathcal{N}_p(0, \boldsymbol{\Sigma})$, and to approximate tight LD blocks, the covariance matrix was constructed by aggregating 8 highly correlated 3x3 blocks $\Sigma = \{\sigma_{ij}\}_{i,j=1}^{3}$, $\sigma_{ij} = 0.95$ if $i \neq j$ and $\sigma_{ii} = 1$. 8 causal variants were distributed across the 8 tight LD blocks, and the remaining variants are designated to be non-causal. Variants have effect sizes of $\pm 1.5$ on the phenotype $\boldsymbol{y}$, which was simulated following the linear model: $\boldsymbol{y} \sim \mathcal{N}_n(\boldsymbol{X\beta}, \boldsymbol{I})$. Bai et al. (2021)

##### S3.1.2 Data pre-processing and Quality Control of real UK Biobank data

The Data pre-processing and Quality Control (QC) pipeline on a sample of 337 205 unrelated White British samples in the UK Biobank (Bycroft et al., 2018) yielded a set of 1 093 308 high quality SNPs for the real data analysis.

The genotype data, which represents the features used for prediction, was extracted by applying standard quality control filters along the sample and variant dimensions.

We now detail the standard quality control filters along the sample and variant dimensions for the genotype data, which represents the features used for prediction. For the samples, in addition to filtering to unrelated White British individuals, we excluded samples with sex chromosome aneuploidy or missing genotype call rate exceeding 5%. The genetic variants were filtered to retain variants in the HapMap3 reference panel (Altshuler et al., 2010), while excluding markers with minor allele count (MAC) <5, minor allele frequency (MAF) <0.1%, imputation score <0.3, Hardy-Weinberg Equilibrium p-value $<10^{-10}$, or genotype missingness rate exceeding 5%. Furthermore, we removed variants with duplicated `rsIDs`, ambiguous strand, or those found in long-range LD regions (Zabad et al., 2023). This resulted in a set of 1 093 308 high quality SNPs that we used in subsequent analyses. Most of these filtering steps were done with `plink2` (Chang et al., 2015).

For the complex traits, each phenotype was pre-processed by removing outliers (z-score >5), and then within each sex separately, the phenotype was adjusted for age, sex, and 10 Principal Components (PCs) of the genotype matrix. As a final step, we applied the Rank-based Inverse Normal Transform (RINT) to ensure that the distribution of the phenotype is approximately Gaussian.

#### S3.2 Derivations of main SSL equations

**Objective function.** We maximize

$$\begin{aligned}\log p(\boldsymbol{\beta}|\boldsymbol{y},\boldsymbol{X},\theta) &= \log p(\boldsymbol{y}|\boldsymbol{X},\boldsymbol{\beta},\sigma^2) + p(\boldsymbol{\beta}|\theta) + \text{const}\\ &= -\frac{1}{2\sigma^2}||\boldsymbol{y}-\boldsymbol{X}\boldsymbol{\beta}||_2^2 + \sum_{j=1}^{p}\log p(\beta_j|\theta) + \text{const}\end{aligned}$$

First, prior can be marginalized to $p(\beta_j \mid \theta)$, which is

$$\begin{aligned}p(\beta_j \mid \theta) &= \sum_{\gamma_j\in\{0,1\}} p(\beta_j,\gamma_j \mid \theta)\\ &= \sum_{\gamma_j\in\{0,1\}} p(\beta_j \mid \gamma_j)p(\gamma_j \mid \theta)\\ &= \sum_{\gamma_j\in\{0,1\}} \psi(\beta_j \mid \lambda_{\gamma_j})\theta^{\gamma_j}(1-\theta)^{1-\gamma_j}\\ &= (1-\theta)\psi(\beta_j \mid \lambda_0) + \theta\psi(\beta_j \mid \lambda_1)\end{aligned}$$

and

$$\begin{aligned}\log p(\beta_j \mid \theta) &= -\lambda_1|\beta_j| + \log\left(\frac{(1-\theta)\,\lambda_0}{\theta\lambda_1}e^{-(\lambda_0-\lambda_1)|\beta_j|}+1\right) + \text{const}\\ &= -\lambda_1|\beta_j| + \log\left(\frac{1}{p^{\star}_{\theta}(\beta_j)}\right) + \text{const}\\ &= -\lambda_1|\beta_j| + \log\left(\frac{p^{\star}_{\theta}(0)}{p^{\star}_{\theta}(\beta_j)}\right) + \text{const}\\ &= \text{pen}(\beta_j \mid \theta) + \text{const}\end{aligned}$$

which gives

$$\log p(\boldsymbol{\beta}|\boldsymbol{y},\boldsymbol{X},\theta) = -\frac{1}{2\sigma^2}||\boldsymbol{y}-\boldsymbol{X}\boldsymbol{\beta}||_2^2 + \sum_{j=1}^{p}\text{pen}(\beta_j \mid \theta) + \text{const} \tag{S1}$$

##### S3.2.1 Update equations in terms of summary statistics

In the following section, with the assumption thhat both the genotype matrix $\boldsymbol{X}$ and phenotype vector $\boldsymbol{y}$ have been standardized column-wise for unit variance and zero mean for GWAS summary statistics, the following equations hold:

$$\begin{aligned}X_j^{\top}X_j &= n\\ X_j^{\top}X_m &= nR_{jm}\\ X_j^{T}\boldsymbol{y} &= n\hat{\beta}_j\\ \boldsymbol{y}^{T}\boldsymbol{y} &= n\end{aligned}$$

where $n$ is the GWAS sample size, $\boldsymbol{R}$ is the $p \times p$ Linkage Disequilibrium (LD) matrix, and $\hat{\beta}_j$ is the standardized marginal GWAS effect size for SNP $j$. Then,

$$\begin{aligned}||\boldsymbol{y} - \boldsymbol{X}\boldsymbol{\beta}||_2^2 &= \boldsymbol{y}^\top \boldsymbol{y} - 2\boldsymbol{y}^\top \boldsymbol{X}\boldsymbol{\beta} + \boldsymbol{\beta}^\top \boldsymbol{X}^\top \boldsymbol{X}\boldsymbol{\beta} \\ &= N(1 - 2\sum_{j=1}^{p} \hat{\beta}_j \beta_j + 2\sum_{j=1}^{p}\sum_{m<j} R_{jm}\beta_j\beta_m + \sum_{j=1}^{p} \beta_j^2)\end{aligned}$$

which can then convert the objective (Eq. S1)

$$L(\boldsymbol{\beta}) = -\frac{n}{2\sigma^2}(1 - 2\sum_{j=1}^{p} \hat{\beta}_j \beta_j + 2\sum_{j=1}^{p}\sum_{m<j} R_{jm}\beta_j\beta_m + \sum_{j=1}^{p} \beta_j^2) - (n+2)\log\sigma + \sum_{j=1}^{p} \text{pen}(\beta_j \mid \theta_j) \tag{S2}$$

and sigma update equation S4 to be in summary statistics. $z_j$ can be computed in terms of summary statistics by

$$\begin{aligned}z_j &= X_j^\top(\boldsymbol{y} - \sum_{k \neq j} X_k \beta_k) \\ &= N(\hat{\beta}_j - \sum_{k \neq j} R_{jk}\beta_k)\end{aligned} \tag{S3}$$

#### S3.3 Unknown variance case

$\sigma^{2(k)}$ can be updated in terms of summary statistics by

$$\sigma^2 = \frac{n}{n+2}(1 - 2\sum_{j=1}^{p} \hat{\beta}_j \beta_j + 2\sum_{j=1}^{p}\sum_{m<j} R_{jm}\beta_j\beta_m + \sum_{j=1}^{p} \beta_j^2) \tag{S4}$$

and the main coordinate ascent and dynamic posterior exploration algorithms (S1, S2) can be adopted to the unknown variance for summary statistics. $\Delta, \beta_j, \theta$ are updated the same way as the fixed variance case in the main text. Since many suboptimal modes exist earlier on in the ladder, (Ročková and George, 2018) recommends updating $\sigma^2$ when the previous $\lambda_0^l$ fit converges in less than 100 iterations. Though, it is suggested to estimate variance in cases where the data isn't standardized to have unit variance (Moran et al., 2019). We describe the effects of the unknown variance case on the simulated UK Biobank data in section S4.3.2.

**Algorithm S1** SSLPRS Coordinate Ascent Unknown Variance.

Input: $\lambda_0, \lambda_1, a, b$
Initialize of $\boldsymbol{\beta}^{(0)} = \mathbf{0}_p, \theta^{(0)} = 0.5, \sigma^{2(0)}, M = 10$
**for** $k$ in $1 \dots k_{max}$ **do**
    Update $\Delta^{(k)} = \begin{cases} \sqrt{2n\sigma^2 \log[1/p^*_\theta(0)]} + \sigma^2\lambda_1 & \text{if } g_\theta(0) > 0 \\ \sigma^2\lambda^*_\theta(0) & \text{otherwise} \end{cases}$
    $p^*_\theta(\beta_j) = \frac{1}{1+\frac{1-\theta}{\theta}\frac{\lambda_0}{\lambda_1}e^{-|\beta_j|(\lambda_0-\lambda_1)}}$,
    $\lambda^*_\theta(\beta_j) = \lambda_1 p^*_\theta(\beta_j) + \lambda_0\big[1 - p^*_\theta(\beta_j)\big]$,
    $g_\theta(\beta_j) = \big[\lambda^*_\theta(\beta_j) - \lambda_1\big]^2 + \frac{2n}{\sigma^2}\log\big[p^*_\theta(\beta_j)\big]$.
    **for** $j$ in $1 \dots p$ **do**
        Compute $z_j = n(\hat{\beta}_j - \Sigma_{k\neq j} R_{jk}\beta_k)$
        Update each $\beta_j^{(k)} \leftarrow \frac{1}{n(1+\lambda_{\min})}\big[|z_j| - \lambda^*_\theta(\beta_j^{(k-1)})\big]_+ \operatorname{sign}(z_j)\, \mathbb{I}(|z_j| > \Delta^{(k)})$
        **if** M updates of $\beta_j^{(k)}$ have occurred **then**
            Update $\theta^{(k)} = \frac{a+\hat{p}_\gamma}{a+b+p}$, $\hat{p}_\gamma = \#\{\beta_j^{(k)} \neq 0\}$
            Update $\sigma^{2(k)} = \frac{n}{n+2}(1 - 2\sum_{j=1}^p \hat{\beta}_j\beta_j^{(k)} + 2\sum_{j=1}^p \sum_{m<j} R_{jm}\beta_j^{(k)}\beta_m^{(k)} + \sum_{j=1}^p(\beta_j^{(k)})^2)$
        **end if**
    **end for**
    **if** $\forall j \ |\beta_j^{(k)} - \beta_j^{(k-1)}| \leq \epsilon_{conv}$ **then**
        break
    **end if**
**end for**

**Algorithm S2** SSLPRS Warm-start. Unknown Variance.

Input: $\lambda_1, a, b$ and ladder of $\lambda_0 = \{\lambda_0^1, \dots, \lambda_0^L\}$
Initialize of $\boldsymbol{\beta}^{(0)} = \mathbf{0}_p, \theta^{(0)} = 0.5, \sigma^{2(0)}, M = 10$
**for** $l$ in $1, \dots, L$ **do**
    Initialize $\lambda_0 = \lambda_0^l, \boldsymbol{\beta} = \boldsymbol{\beta}^{(l-1)}, \sigma^2 = \sigma^{2(l-1)}, \theta = \theta^{(l-1)}$
    Initialize $\sigma^2_{\text{update}}$ = False
    **if** $l \neq 1$ and $k^{(l-1)} < 100$ **then**
        $\sigma^2_{\text{update}}$ = True
    **end if**

    Run coordinate ascent with Algorithm S1.

    $(\boldsymbol{\beta}^{(l)} = \boldsymbol{\beta}, \sigma^{2(l)} = \sigma^2, \theta^{(l)} = \theta)$
**end for**

#### S3.4 Derivations of alpha heritability model *SSLPRS-Alpha*

Studies have shown that there is a non-linear relationship between the minor allele frequency (MAF) of a genetic variant and its effect size (Speed et al., 2020; Gazal et al., 2017; Zeng et al., 2021). This observation has motivated per-SNP heritability models, where the expected contribution of each SNP is scaled by its allele frequency $p_j$. To incorporate this dependence into a penalized regression framework, we adapt the alpha prior heritability model to the SSL model, and derive the *SSLPRS-Alpha* model, where:

$$\alpha \text{ model: } \mathbb{E}[h_j^2] \propto v_j^{1+\alpha}, \quad \mathrm{Var}(\beta_j^{(\text{allele})}) \propto v_j^{\alpha}$$

Here, $v_j = \mathrm{Var}(G_j) = 2p_j(1-p_j)$ is the genotypic variance of variant $j$ and $\beta_j^{(\text{allele})}$ is the un-scaled per-allele effect size. Upon standardizing $G$ column-wise to have zero mean and unit variance for GWAS summary statistics, we obtain the standardized genotype matrix $X$. The per-SNP variance for of the effect size $\beta_j$ for $X$ becomes:

$$\begin{aligned}\mathrm{Var}(\beta_j) &= \mathrm{Var}(\sqrt{v_j}\beta_j^{(\text{allele})}) \\ &\propto v_j \mathrm{Var}(\beta_j^{(\text{allele})}) \\ &= v_j v_j^{\alpha} \\ &= v_j^{\alpha+1}\end{aligned}$$

Reparameterizing the Spike-and-Slab Laplace (SSL) penalties $\lambda_0$ and $\lambda_1$ to match $\mathrm{Var}(\beta_j)$ gives:

$$\begin{aligned}\mathrm{Var}(\beta_j) &= \frac{2}{\lambda_{1,j}^2} \propto v_j^{\alpha+1} & \qquad \mathrm{Var}(\beta_j) &= \frac{2}{\lambda_{0,j}^2} \propto v_j^{\alpha+1} \\ \Rightarrow \lambda_{1,j} &\propto v_j^{-(\alpha+1)/2} & \Rightarrow \lambda_{0,j} &\propto v_j^{-(\alpha+1)/2}\end{aligned}$$

Thus, the Laplace densities become SNP specific, where the spike and slab penalties for SNP j is defined as $\lambda_{0,j} = \lambda_0 v_j^{-(\alpha+1)/2}$ and $\lambda_{1,j} = \lambda_1 v_j^{-(\alpha+1)/2}$ respectively, where $\lambda_0$ and $\lambda_1$ are the uniform spike and slab penalties in the original *SSLPRS* model. The *SSLPRS-Alpha* model is also available in the `penprs` package under the `SSLAlpha` class.

To evaluate the model performance of the alpha heritability SSL model, we compare the grid-searched *SSLPRS-Alpha-GS*, with the default $\alpha = -0.25$ model (Speed et al., 2020; Zeng et al., 2021), to the grid-searched *SSLPRS-GS* across the nine real phenotypes from the UK Biobank in Figure S8. Although the *SSLPRS-Alpha* model may not have consistent or substantial gains, similar to the results illustrated in the alpha modeled version of *VIPRS* (Zabad et al., 2023), it may become more beneficial when integrated with other omics modalities and other factors that capture more non-linear dependencies.

#### S3.5 Stability of model training

In the analyses conducted in this study, *SSLPRS* utilized Linkage-Disequilibrium (LD) matrices from a reference panel of 342 446 unrelated European samples in the UK Biobank (Bycroft et al., 2018). For computational efficiency and stable training, we used sparse block-diagonal LD estimators, with blocks defined by `LDetect` software (Berisa and Pickrell, 2016). However, block-diagonal LD matrices may still have negative eigenvalues, which could cause optimization algorithms to pursue directions aligned with them, leading effect size estimates to explode and diverge (Zabad et al., 2025).

To ensure training stability, we introduce an additional $\lambda_{\min}$ penalty following the approach outlined in (Zabad et al., 2025; Choi and Tibshirani, 2013). Specifically, given the original LD matrix $\tilde{\boldsymbol{R}}$, we use $\tilde{\boldsymbol{R}} + |\lambda_{\min}|I$ as the LD, where $I$ is the identity matrix and $|\lambda_{\min}|$ is the absolute value of the minimum eigenvalue of $\mathbf{R}$ (i.e., the magnitude of the most negative eigenvalue) to ensure $\tilde{\boldsymbol{R}}$ is positive semi-definite. This approach is similar to *Lassosum* (Mak et al., 2017; Privé et al., 2022), where the authors introduced the parameter $s \in [0, 1]$ to scale the original LD as $\tilde{\boldsymbol{R}} = (1 - s)\tilde{\boldsymbol{R}} + sI$ to ensure stability. However, *Lassosum* treats $s$ as a hyperparameter to perform grid search over, with default grid of $[0.2, 0.5, 0.9, 1]$. This additional penalty makes both models more similar to an elastic-net penalization technique rather than a pure Lasso one, as noted by Choi and Tibshirani (2013) and the original authors of the *Lassosum* method. However, in our case, instead of treating $\lambda_{\min}$ as a hyperparameter, we derive it directly from spectral properties of the LD matrix.

In addition, we note that the spectral properties of LD matrices can be affected by many choices, such as sparsification mask or compression scheme. For example, using less aggressive quantization schemes, such as `int16`, in place of the default `int8` used for LD matrices from (Zabad et al., 2025), results in substantially lower $\lambda_{\min}$ values, thereby reducing shrinkage and bringing *LASSO* closer to a standard Lasso solution. This has minimal to no measurable impact on model performance as noted in section S3.6 while improving model training stability, but at the cost of increased memory usage. A close look at the effects of $\lambda_{\min}$ can be found in section S4.3.1, where we compare the model stability across different $\lambda_{\min}$ values. This distinction is a key factor underlying the differences in the predictive performance on the real phenotypes (Figure 3, main text), which we discuss in greater details in section S4.2 together with other differences.

#### S3.6 Software and runtime optimizations

*SSLPRS* was implemented mainly in `Python`, except expensive computations in the coordinate ascent algorithm, which was implemented using the compiled language via the `Cython` interface for compatibility and optimal efficiency. We also employed model specific optimization and similar optimization strategies used in *VIPRS* (Zabad et al., 2023, 2025), including $q$-factors to access the LD matrix only once per iteration, which we describe in detail below. We also introduced parallelism in the form of multi-threading across SNP updates in the main coordinate ascent loop (Algorithm S1), which improves overall inference time by multi folds. To reduce overall model peak memory usage, we introduced a low memory mode, which reduces storage requirements by more than a factor of two by storing only the upper-triangular part of the symmetric LD matrix. Further memory optimizations were achieved through floating point quantization to integer representation, minimizing memory footprint for each entry of the LD while having a minimal impact on performance (Zabad et al., 2025). In addition, quantities like $\hat{p}_\gamma$ are updated on the fly to eliminate inefficient re-computations.

A major model specific efficiency optimization we introduced leverages *SSLPRS*' and *LASSO*'s ability to produce exactly sparse effect size estimates. For the subset of $\beta_j$ coefficients that are identical after the update, we skip expensive computations such as $q$-factor updates that wouldn't have had any effects on the model parameters. This greatly speeds up the time per iteration of the coordinate ascent algorithm, especially for large $\lambda_1$ penalty values, where many of the effect sizes are shrunk to zero.

##### S3.6.1 $q$-factor optimization

To minimize the number of LD accesses per iteration, which can induce a bottleneck in the main coordinate ascent updates, we introduce a $q$-factor quantity for each SNP in a similar fashion in (Zabad et al., 2023). The $q$-factor, for a SNP $j$ is defined as

$$q_j = \sum_{k \neq j} \beta_k R_{jk}$$

and when $\beta_j$ is updated to a new estimate $\beta_j^{(new)}$, a partial update to the q factors of SNPs belonging to the neighborhood of SNP $j$ can be done by:

$$q_k \leftarrow q_k + R_{jk}(\beta_j^{(new)} - \beta_j^{(old)}) \quad \forall k \neq j$$

When $\beta_j^{(new)} = \beta_j^{(old)}$, then the q factor for SNP $j$ remains the same. Thus, the $q$-factor update can be skipped for all neighbors of SNP $j$, which is the main optimization we perform that takes advantage of *SSLPRS*' exactly sparse estimates. Previously, the LD-matrix $R$ requires access in 3 separate updates and computations of the algorithm, specifically Eq. S3, S4 and S2, which has now been reduced to one with the following equations utilizing $q$-factors:

$$z_j = N(\hat{\beta}_j - q_j)$$

$$\sigma^2 = \frac{N}{N+2}(1 - 2\sum_{j=1}^{p} \hat{\beta}_j \beta_j + \sum_{j=1}^{p} \beta_j q_j + \sum_{j=1}^{p} \beta_j^2)$$

and the objective can be updated as

$$L(\boldsymbol{\beta}) = -\frac{N}{2\sigma^2}(1 - 2\sum_{j=1}^{p} \hat{\beta}_j \beta_j + \sum_{j=1}^{p} \beta_j q_j + \sum_{j=1}^{p} \beta_j^2) - (n+2)\log\sigma + \sum_{j=1}^{p} \mathrm{pen}(\beta_j|\theta_j) \tag{S5}$$

### S4 Supplemental Results

#### S4.1 Variable selection behavior across warm-start ladder on simulated setting of UK Biobank phenotype data

To illustrate the variable selection behavior of *SSLPRS* and *LASSO* across the $\lambda_0$ ladder, we focus on chromosome 1 of the balanced $h^2 = 0.3, \theta = 0.001$ setting. As shown in Figure S1, a tradeoff between the precision and recall emerges as $\lambda_0$ increases. In the earlier steps of the ladder, where the difference between $\lambda_0$ and $\lambda_1$ is marginal, *SSLPRS* remains effectively in the LASSO regime — yielding good recall, but suboptimal precision due to over-selection (Zhao and Yu, 2006). As *SSLPRS* warm-starts through the ladder, negligible coefficients are progressively shrunk to zero, and significant variants are retained with increasing confidence. This leads to a substantial gain in precision, but a decrease in recall. While *LASSO* exhibits a similar tradeoff, the absence of a stabilizing slab results in a sharp deterioration of recall without a commensurate increase in precision. Notably, the $\lambda_0$ selected by *SSLPRS-GS* and *LASSO*, which is coincidentally the same in this case, occurs earlier in the ladder, optimizing pseudo-$R^2$ but may not yield the best variable selection in terms of Matthew Correlation Coefficient (MCC). As a result, *SSLPRS*, which warm-starts to the end of the ladder, achieves superior overall performance with respect to MCC compared to baseline models.

#### S4.2 Comparisons between *LASSO* and *Lassosum*

As part of our `penprs` software, we implemented our own version of the *LASSO* model. *LASSO* naturally takes advantage of the computational improvements outlined in Section S3.6, making it highly efficient among the methods as seen in S14. In the analyses for predictive performance on the real phenotypes, we included an external model, *Lassosum* (Mak et al., 2017), which is also based on the Lasso prior. However, variations in LD reference panel, penalty ladder formulation, and implementation details can lead to differences in predictive performance across phenotypes. For instance, *LASSO* performs better in some traits (e.g., HDL, LDL) whereas *Lassosum* performs better in others (e.g., BMI, FEV1), as illustrated in Figure 3 of the main text. We examine the variations in detail in the subsections below.

##### S4.2.1 Differences in LD scaling and reference panel

The performance difference between *LASSO* and *Lassosum* arises mainly due to how each method represents and scales the LD matrix during training. As described in detail in section S3.5, *LASSO* uses a $\lambda_{min}$ penalty, with the quantity determined by the spectral properties of the LD matrix. On the other hand, *Lassosum* implements a grid search for the hyperparameter $s \in [0.2, 0.5, 0.9, 1]$, which is a strong penalty that causes *Lassosum* to behave more similar to elastic-net rather than *Lasso*. As noted in S3.5, LD matrices used in the analyses of this study require very low magnitudes of $\lambda_{min}$ for stabilized training, introducing only a negligible degree of ridge penalization for *LASSO*. Consequently, in Figure S12, when comparing *LASSO* to *Lassosum* in the earlier steps of the $\lambda$ penalty ladder, where most effect size estimates are non-zero, the estimates deviate more for higher values of $s$, rendering *Lassosum* increasingly similar to an elastic-net model. Specifically, if *Lassosum* is fitted with an extremely small $s = 10^{-8}$ value, where it behaves like standard Lasso, the effect size estimates of *LASSO* and *Lassosum* are nearly perfectly correlated across the $\lambda$ penalty ladder. However, the smallest default value of $s$ considered by *Lassosum* is 0.2, which already introduces a moderate ridge penalty and becomes more elastic-net like, resulting in differences in effect size estimates. This becomes more apparent for the larger $s = 0.5$ case, and in practice, $s$ can be selected to be as large as $s = 1$, driving *Lassosum* far from the standard Lasso, thereby resulting in large differences when compared to *LASSO*.

##### S4.2.2 Methodological nuances and implementation details

In addition to the differences in LD scaling between *LASSO* and *Lassosum*, which account for the primary differences in performance, there are other methodological and implementation differences that contribute to more subtle variations in performance. First, *Lassosum* constructs its penalty ladder in a genome-wide manner, fitting all chromosomes under the same ladder and selecting the single best genome-wide performing $\lambda$ based on validation performance. In comparison, *LASSO* formulates the penalty ladder, through the data specific $\lambda_{max}$, the minimum penalty beyond which all negligible coefficients would be set to zero (Friedman et al., 2010), separately for each chromosome, and selects the optimal $\lambda$ on a per-chromosome basis. Furthermore, in combination with summary statistics, *Lassosum* requires a genotype reference panel (e.g., 1000 Genomes, UK Biobank) of individual-level data to implement the update equations, whereas *LASSO* uses user-supplied LD matrix.

#### S4.3 Ablation of hyperparameters

To evaluate the robustness of *SSLPRS* under varying hyperparameter settings, we conducted a series of ablation studies on its key hyperparameters to demonstrate the effects on training, variable selection, and predictive performance. Specifically, other than the grid-searched spike penalty $\lambda_0$ across the warm-start ladder, we investigated: (i) the effects of $\lambda_{min}$ on model training stability and inference (ii) the sensitivity of variance $\sigma^2$, (iii) the selection of the slab penalty $\lambda_1$, (iv) the prior mean on the mixing proportion $\theta$, and (iv), different $\lambda_0$ penalty ladder lengths. In addition, we evaluated the sensitivity of *VIPRS* on different Posterior Inclusion Probability (PIP) thresholding values for variable selection and SNP significance replication performance.

In addition, based on these analyses, users can leverage the results to better understand which hyperparameters are most effective for specific tasks or scenarios. Accordingly, they may adjust the hyperparameter configurations to optimize training stability, inference accuracy (e.g., pseudo-Pearson-$R^2$), or specific variable selection metrics such as precision, recall, or the overall MCC.

##### S4.3.1 Effects of $\lambda_{min}$ on model training stability and inference

As described in detail in section S3.5, $\lambda_{min}$ is a important data-derived quantity that stabilizes model training, preventing the model from pursuing optimization directions that would cause diverging effect size estimates. In Figure S11, we conducted an ablation to illustrate the effects and importance of $\lambda_{min}$ on a range of values using `int8` quantized LD matrices, as they require larger $\lambda_{min}$ values to stabilize and therefore highlight the differences more clearly.

In Figure S11a, when trained on $\lambda_{min} = 0$ or $\lambda_{min} = 0.1$ values, the effect size estimates diverge over iterations, with the instability occurring more rapidly with $\lambda_{min} = 0$. As the magnitude of $\lambda_{min}$ increases toward the value derived from the spectral properties of the LD matrix, training becomes more stable, and larger values results in faster convergence as there's more regularization. Although $\lambda_{min} = 0.15$ remains stable in this particular case, we still recommend using the LD-derived $\lambda_{min}$ to ensure the LD matrix is positive semi-definite. In this case, it's the starred $\lambda_{min} = 0.22$ value. As illustrated in S11b, if the model converges, *SSLPRS* is robust to the choice of $\lambda_{min}$ in terms of predictive performance, with pseudo-Pearson $R^2$ differing only slightly. As the $\lambda_{min} = 0.22$

value derived from the LD matrix achieves performance that is essentially indistinguishable from the top performance values, in general, *SSLPRS* uses it as the default $\lambda_{min}$ value without any hyperparameter-search.

##### S4.3.2 Sensitivity of variance $\sigma^2$ and the unknown variance case

Estimating variance $\sigma^2$, as described in section S3.3 instead of fixing it could prove to be beneficial in cases where the data isn't standardized, as illustrated in (Moran et al., 2019). However, when the data is assumed to be scaled to have unit variance, such as the data used in the real and simulated analyses, estimating $\sigma^2$ showed little differences compared to the fixed variance $\sigma^2 = 1$ case (Figure S7). While *SSLPRS-GS* showed modest gains in variable selection performance, the best performing default variant *SSLPRS* exhibited reduced performance under the unknown variance case. We also found that in terms of pseudo-Pearson-$R^2$ predictive performance, *SSLPRS-GS* is robust in the selection of $\sigma^2$ in the fixed case when trained on standardized, unit variance data. In additional, $\sigma^2$ updates significantly increase computational costs, with up to a 6 fold increase in total runtime for *SSLPRS*. Thus, we decided to fix the variance $\sigma^2 = 1$ for the analyses conducted in this study.

##### S4.3.3 Selection of $\lambda_1$ through the scale hyperparameter $\epsilon$

As noted in the main text, by default, we set the slab penalty $\lambda_1$ relative to the $\lambda_{max}$ penalty, the minimum penalty beyond which all negligible coefficients would be set to zero (Friedman et al., 2010), and to ensure the slab is sufficiently diffuse, $\lambda_1 = \epsilon\lambda_{max}$, where by default, the scale hyperparameter is set to $\epsilon = 10^{-3}$. This scaling also preserves the ratio between $\lambda_1$ and the spike penalty $\lambda_0$ fixed across different chromosome and dataset training, since both are defined relative to $\lambda_{max}$. The effects of $\epsilon$ are illustrated in Table $S2$, where we conduct an ablation over a range of $\epsilon \in [0.0001, 0.001, 0.01, 0.1, 0.25]$ values, comparing variable selection and predictive performance. In all values except $\epsilon = 0.25$, which the worst performing setting, the performance of *SSLPRS* and *SSLPRS-GS* is similar, and for the default $\epsilon = 10^{-3} = 0.001$ choice, the respective *SSLPRS* variants achieves the best performance in the task it is optimized for (i.e., pseudo-Pearson-$R^2$ for *SSLPRS-GS* and overall MCC for *SSLPRS*). Thus, we keep $\epsilon = 10^{-3}$ for the slab penalty $\lambda_1 = \epsilon\lambda_{max}$ to resemble a diffuse slab relative to the spike.

##### S4.3.4 Effects of prior mean on mixing proportion $\theta$

An important variable selection quantity is the prior mean of the mixing proportion $\theta$, and since $\theta \sim Beta(a, b)$, $\mathbb{E}(\theta) = \frac{a}{a+b}$, where $a$ and $b$ are hyperparameters. To tune the prior mean, we keep $b = p$ fixed and tune $a$ to be $a = \frac{x}{1-x}b$, where $x$ represents the desired mixing proportion value. To illustrate the effects of different prior mean values, we conduct an ablation over $\mathbb{E}(\theta) \in [0.01, 0.05, 0.10, 0.15, 0.2]$ in Table S2. Across the different values, the pseudo-Pearson-$R^2$ is relatively the same for the prediction focused *SSLPRS-GS*. In terms of the variable selection focused *SSLPRS*, we observe a trade-off between precision and recall. Lower prior mean values $\mathbb{E}(\theta)$ yield higher precision at the cost of sensitivity, as fewer variants are selected, but the model is more confident about the selection. In contrast, higher $\mathbb{E}(\theta)$ values lead to increased recall, since more variants are selected when the mixing proportion is larger. Overall, the most balanced setting is $\mathbb{E}(\theta) = 0.05$, which we set as the default, where the MCC is the best among the values for *SSLPRS*.

##### S4.3.5 Effects of spike penalty $\lambda_0$ warm-start ladder length

By default, we perform warm-start fit across a log scaled $\lambda_0$ ladder of 20 steps. To assess the robustness of *SSLPRS* on different ladder lengths, we conduct an ablation in Table S2 on ladder lengths of $[10, 20, 30]$. Both *SSLPRS* and *SSLPRS-GS* perform the best in the metric they are optimized for (i.e., pseudo-Pearson-$R^2$ for *SSLPRS-GS* and overall MCC for *SSLPRS*. We therefore adopt a balanced 20 step $\lambda_0$ ladder for a fine grid with enough values that can effectively explore the parameter space without being too coarse like a 10-step grid or being too fine, such as a 30-step ladder where it might select values that overfits to noise. Thus, we choose a $\lambda_0$ ladder length of 20 as the default.

##### S4.3.6 Sensitivity of PIP thresholds for *VIPRS* variable selection

We assessed the sensitivity of VIPRS to different posterior inclusion probability (PIP) thresholds for variable selection, comparing values of 0.3, 0.5 (median rule), 0.7, 0.9. As illustrated in Table S4, the number of selected variants decreased as the PIP threshold increased, selecting variants that the model thinks are more likely to be causal. At the same time, the Benjamini–Hochberg (BH) (Benjamini and Hochberg, 2018) FDR ($\alpha = 0.05$) controlled replication rate increased at the cost of decreased FDR-controlled replication count, based on the significance indicated in the independent GLGC meta-analysis (Graham et al., 2021). Specifically, under the BH procedure, the selected variants' p-values according to the replication study are first ranked in ascending order, and the largest $k$ is identified such that $p_{(k)} \leq \frac{k}{m}\alpha$, where $p_{(k)}$ is the $k$-th smallest p-value, $m$ in this case, is the number of selected variants by VIPRS based on the PIP thresholding, and $\alpha = 0.05$ is the desired FDR level.

In terms of variable selection, performance is quite similar between the PIP threshold values. This is likely due to a characteristic of variational Bayesian methods for variable selection in regression, which tend to push PIP values into saturated bins near the high and low distributions of PIP, leaving variants in the intermediate range (Carbonetto and Stephens, 2012), thereby making the model not too sensitive to the threshold choice. Though, a slight tradeoff is apparent: higher thresholds have higher precision, as the model is more confident about the significance of the selected variants, while lower thresholds yield higher recall, as a larger set of variants is retrained. Overall, the balanced threshold 0.5 has the best MCC compared to the other values. Therefore, we decided to use the median rule $PIP > 0.5$ (Ishwaran and Rao, 2005), to have a good balance between number of FDR-controlled replication count and replication rate and a good balance of precision and recall in variable selection.

#### S4.4 Transferability to non-European minority populations

To assess the transferability of model performance to non-European minority populations, we first trained the models using summary statistics of the White British cohort and the European LD reference panel. Then model effect size estimates were transferred to minority populations: Italy, China, India, and Nigeria and evaluated using their respective LD reference panel: European (EUR), East Asian (EAS), Central/South Asian (CSA), and African (AFR) to evaluate predictive performance across nine quantitative phenotypes using pseudo-Pearson-$R^2$ on the minority summary statistics, as illustrated in panels of Figure S10.

Overall, prediction accuracy in terms of pseudo-Pearson-$R^2$ declined as genetic distance increased with respect to the White British population. The relative performance between the models remained largely the same when evaluated on a minority population across the nine real quantitative phenotypes, suggesting that ancestry, via differences in allele frequencies and LD, rather than model algorithm and formulation, is the primary determinant of portability. However, since *SSLPRS* was not explicitly optimized for cross-ancestry transfer, it tended to underperform slightly in some minority ancestry groups such as China (EAS) or India (CSA) relative to the other models with respect to the White British cohort. A plausible explanation could be that although *SSLPRS* produces high quality and unbiased sparse effect size estimates in the discovery population, it may lock onto ancestry-specific SNPs calibrated to the EUR LD. As LD patterns and allele frequencies shift in target transfer minority ancestry, the effect size estimates could be overestimated. By contrast, methods such as *LASSO* that have more uniform shrinkage with respect to *SSLPRS* could be more biased in the discovery population, as would be large effect size estimates are shrunk as well, distribute smaller and more conservative weights that are less impacted to LD shifts, yielding a slightly smaller transfer penalty.

#### S4.5 Regression of phenotypes calibration check on PRS quantiles

To evaluate the calibration of the polygenic risk scores (PRS), we regress the phenotype values of individuals against deciles of the predicted PRS. Individuals were stratified into ten equal-size quantile bins, where quantile 1 (Q1) contain the lowest predicted PRS individuals, and quantile 10 (Q10) contain the highest predicted PRS individuals. In Figure S13, we perform the regression calibration on chromosome 22 of the standing height (HEIGHT) phenotype from the UK Biobank and a clear monotonic increase is observed in the mean phenotype values across the PRS quantiles. Specifically, individuals in lower predicted PRS quantiles exhibited lower mean phenotype values (height), whereas those in higher PRS quantiles displayed progressively greater mean values in the regression, indicating a good calibration and meaningful discrimination power of the PRS.

#### S4.6 Computational performance and scalability of *SSLPRS*

Following the enhancements we introduced in Section S3.3, we compared our proposed summary statistics version *SSLPRS* to the individual level, `R`-language based, `SSLASSO` from the original authors Ročková and George (2018) to highlight the computational efficiency and also scalability across an increasing sample size and variant count of *SSLPRS*. Both models were benchmarked on identical, sample size $n$ and variant count $p$ subsets of standing height data from the UK Biobank on chromosome 22. Specifically, we evaluated the models across sample sizes $n \in \{10000, 25000, 50000\}$, and SNP counts $p \in \{5000, 10000, 16296\}$. For each $(n, p)$ combination, we recorded the total inference time and peak memory usage, across the default *SSLPRS* warm-start ladder. As seen in Table S5, across all settings, *SSLPRS* had an average inference time of 5.3 s and peak memory usage of 73 MB, whereas the original individual-level *SSL* required 22.4 min and 4100 MB, respectively. Notably, our summary statistics implementation maintained near-constant inference time as sample size increased, and scaled efficiently with growing SNP counts. In contrast, the individual-level `SSLASSO` exhibits approximately linear scaling of inference time and peak memory usage as sample size or SNP count increases. These results underscore a significant improvement in computational efficiency and scalability through summary statistics and optimizations. A comprehensive comparison between *SSLPRS* and the baseline summary statistics models regarding computational performance are shown in Figure S14.
